# What is the most effective method of delivering Making Every Contact Count training? A rapid review

**DOI:** 10.1101/2024.10.18.24315722

**Authors:** Leona Batten, Greg Hammond, Clare England, David Jarrom, Elizabeth Gillen, Jacob Rees Davies, Rhiannon Tudor Edwards, Adrian Edwards, Alison Cooper, Ruth Lewis

## Abstract

The Making Every Contact Count MECC programme encourages staff to have opportunistic brief behaviour change conversations with service users. It uses the day-to-day interactions that healthcare professionals, or those within other organisations, including the not-for-profit sector have with people, to support them in making positive changes to their physical and mental health, and wellbeing. The aim of this review is to assess which elements or types of MECC training, or comparable interventions, are most effective and preferred by those who would implement MECC in practice.

The review included evidence available up until June 2024. 11 studies were included. These all focused on healthcare organisations and included health care or public health professionals, with two also including trainees who worked in a local authority.

There was consistent evidence that training increased both trainee confidence and use of MECC-related techniques immediately following training. There was some evidence that despite a slight reduction, these improvements were at least maintained up to one year later. There was no evidence on the longer-term effect, other than an indication that refresher training would be appreciated. There was also no evidence assessing whether improvements in trainee confidence and competence had any impact on service user behaviour change and outcomes. There was an indication that face-to-face training was preferred to online training.

Barriers to MECC training attendance included a feeling that there was not enough time, and a lack of managerial support. Barriers to MECC utilisation included a feeling that there was not enough time, a lack of organisational and managerial support, a fear of upsetting patients, and a lack of awareness of downstream support services to refer service users to following healthy behaviour conversations. The evidence indicated that barriers to MECC training and utilisation could be overcome via provision of information about downstream support services, and improved organisational and managerial support for both attendance at MECC training and its use in practice. Further research is needed. This should include research into the impact from MECC on patient behaviour and eventual outcomes, and how these change following training.

**Funding statement:** The authors and their Institutions were funded for this work by the Health and Care Research Wales Evidence Centre, itself funded by Health and Care Research Wales on behalf of Welsh Government

**EXECUTIVE SUMMARY:** *What is a Rapid Review?:* Our Rapid Reviews (RR) use a variation of the systematic review approach, abbreviating or omitting some components to generate the evidence to inform stakeholders promptly whilst maintaining attention to bias.

*Who is this Rapid Review for?:* The review question was suggested by Cwm Taf Morgannwg University Health Board Public Health Team. The review is intended to inform those responsible for commissioning and leading Making Every Contact Count (MECC) training.

*Background / Aim of Rapid Review:* The MECC programme encourages staff to have opportunistic brief behaviour change conversations with service users. It uses the day-to-day interactions that healthcare professionals, or those within other organisations, including the not-for-profit sector have with people, to support them in making positive changes to their physical and mental health, and wellbeing. The aim of this review is to assess which elements or types of MECC training, or comparable interventions, are most effective and preferred by those who would implement MECC in practice. Findings may be used to inform creation of future training, or to update current offerings, and improve consistency across health boards.

*Results of the Rapid Review:* Recency of the evidence base

- The review included evidence available up until June 2024. Publication dates of included evidence ranged from 2013 to 2023. Extent of the evidence base

- 11 studies were identified for inclusion in this review; all focused on healthcare organisations.
- 10 studies took place in the UK (eight of which were in England, and none in Wales), and one was undertaken in Australia. All studies included health care or public health professionals, with two also including trainees who worked in a local authority.
- Study designs included pre-test/ post-test, post-test only, qualitative study designs (e.g. interviews, surveys), and mixed methods studies. No study included a separate control group.
- Five of the included studies used surveys or questionnaires only, three used interviews only (either with individuals or focus groups), two used a combination of surveys and interviews, and one undertook surveys, interviews and performed observations of participants. Key findings and certainty of the evidence

- There was **consistent evidence that training increased both trainee confidence and use of MECC-related techniques** immediately following training. There was some evidence that despite a slight reduction, these **improvements were at least maintained up to one year later**. There was no evidence on the longer-term effect, other than an indication that **refresher training would be appreciated**. There was also no evidence assessing whether improvements in trainee confidence and competence had any impact on service user behaviour change and outcomes.
- There was an **indication that face-to-face training was preferred to online training**. However, some trainees did seem to prefer online training due to its increased flexibility.
- **Barriers to MECC training attendance** included a feeling that there was **not enough time, and a lack of managerial support**.
- **Barriers to MECC utilisation** included a feeling that there was **not enough time, a lack of organisational and managerial support, a fear of upsetting patients, and a lack of awareness of downstream support services** to refer service users to following healthy behaviour conversations.
- The **evidence indicated** that **barriers to MECC training and utilisation could be overcome** via provision of information about downstream support services, and improved organisational and managerial support for both attendance at MECC training and its use in practice. Policy and Practice Implications

- MECC training can be beneficial in improving trainee confidence and competence in using MECC. However, there are consistently reported barriers both to undertaking training and using MECC in practice, as summarised above. Although this review identified some potential ways of overcoming barriers both for attending training and utilising MECC in practice, the best and most effective ways of overcoming them remain unclear, and many require a widespread culture change.
- The lack of evidence around whether changes to MECC training have an impact on service user behaviour and eventual outcomes, may impact how effective attempted changes to MECC training and workplace culture could be.
- This review indicates that there appears to be a preference for standardised training with some capacity for tailoring to local needs. This should be taken into account when considering any updates to the Welsh MECC training modules.

*Research Implications and Evidence Gaps:*

- There needs to be further research into the impact from MECC on patient behaviour and eventual outcomes, and how these change following training in both the short and long term.
- Research specifically into the MECC training being implemented across Wales, how people interact with and use this, and what they find useful and relevant would also be worthwhile.
- Identification of methods to overcome the specific barriers and enhance the enablers-particularly around encouraging organisational and managerial support for MECC training and utilisation.

*Economic considerations:* The Public Health England MECC evaluation guidance 2020 recommends producing a business case for MECC programmes that includes the costs of delivery and considers the value for money of its implementation. Most policy documents discussing the cost-effectiveness of MECC programmes cite the National Institute for Health and Care Excellence (NICE) public health guidance 2014 (PH49). This guidance however, does not directly refer to MECC programmes, but rather brief behaviour change and signposting interventions.

## 1. BACKGROUND

### 1.1 Who is this review for?

This Rapid Review was conducted as part of the Health and Care Research Wales Evidence Centre Work Programme. The review question was initially proposed and further developed following discussion with Cwm Taf Morgannwg University Health Board (CTMUHB) Public Health Team. The review is intended to inform those responsible for leading, developing and delivering Making Every Contact Count (MECC) training, with the vision of supporting a consistent training offer across Health Boards, resulting in increased use of MECC in day-to-day interactions, with the ultimate aim of improving health outcomes for service users.

### 1.2 Background and purpose of this review

Non-communicable diseases (NCD) such as cardiovascular disease, cancer, diabetes and obesity contribute to a large number of deaths and a high proportion of premature mortality rates, with the population of the UK estimated to have a 10% probability of premature mortality due to NCDs (World Health Organisation 2022). Many elements of these diseases are closely linked to behavioural risk factors, which are impacted by lifestyle and health choices, such as diet, physical exercise and smoking (World Health Organisation 2023). Adopting healthier behaviours can therefore reduce the impact of non-communicable diseases by helping to prevent premature mortality, reduce health inequalities, and help those with long-term conditions manage these better. MECC is an approach to behaviour change which utilises the day-to-day interactions that healthcare professionals, or those within other organisations, including the not-for-profit sector, have with people to support them in making positive changes to their physical and mental health and wellbeing. It is a National (UK) initiative that was introduced in Wales in 2016 (Meade et al. 2023). Within CTMUHB, the MECC initiative focuses on seven key lifestyle behaviours; healthy eating, smoking cessation, being more active, reducing alcohol consumption, looking after wellbeing, keeping up with immunisation, and engagement with screening programmes (Public Health Wales 2024b).

MECC training for healthcare professionals and other organisations is available throughout Wales with national oversight from Public Health Wales. There are 2 levels of MECC training within Wales – Level 1 is a standardised online e-learning training module, open to all individuals (Public Health Wales 2024a). The learning objectives are i) Recognise your role in supporting people to make healthy choices, ii) Understand the key healthy lifestyle messages, iii) Recognise and act on opportunities to have MECC conversations, and iv) Know how to have an effective conversation about healthy lifestyles. Level 1 is mostly aimed at enabling staff to provide service users with brief advice and a healthy chat (Public Health Wales 2017). Level 2 training sits within the seven individual Health Boards across Wales, and the focus and method of delivery varies between them. However generally, level 2 training aims to introduce staff to brief interventions and improve their skills and knowledge in the relevant techniques (Public Health Wales 2017). In CTMUHB, training is delivered either face-to-face or through Microsoft Teams, and is available to anybody who is in a position to hold a healthy lifestyle-based conversation. The learning objectives for staff in CTMUHB are i) To gain understanding of what MECC is, the relevance to your role, and the key lifestyle behaviours it covers, ii) To understand the current picture in CTMUHB through review of the data, iii) To understand what a brief intervention approach is, and the tools that can be used to support healthy conversations, and iv) To gain knowledge of signposting opportunities across CTM. Neither training module is mandatory in Wales.

The variation in the content and delivery of the Level 2 training between Health Boards means there is uncertainty around the best way in which to deliver the training, how this impacts upon trainee reaction, learning and behaviour such as their utilisation of MECC, and how it may then subsequently affect service user outcomes. An evidence review exploring this has the potential to inform both local and national organisational culture, and provide recommendations to support consistency within both the content and mechanisms of training. This, in turn, may support the embedding of MECC into everyday practice for individuals/organisations in a position to hold behaviour change conversations.

The aim of this review is to assess which elements or forms of training for MECC, or similar interventions, are most effective and preferred by those who would implement MECC in practice. Findings can be used to inform creation of future training, or to update current offerings, and improve consistency across Health Boards.

For the purposes of this review, MECC is defined as an opportunistic brief intervention conversation. “Brief Intervention” is defined as a short, structured conversation that offers opportunistic advice, discussion, or negotiation that aims to strengthen a person’s commitment for behaviour change (National Institute for Health and Care Excellence (NICE), 2006). It is a person centred – holistic approach, and is not Motivational Interviewing, goal setting or a reductionist approach.

## 2. RESULTS

### 2.1 Review of MECC training

#### 2.1.1 Overview of the Evidence Base

Full details of study eligibility criteria are described in Section 5, along with the methods used for this review. A detailed summary of the search results and study selection processes are summarised in Section 6.

There were 11 studies identified that answered the review question. An overview of the characteristics of included studies is provided in Table 1. All studies focused on supporting the use MECC by health care professionals (including undergraduate students), with two also reviewing MECC use by local authorities (Chisholm et al. 2020, Nelson et al. 2013). Three studies were pre-post surveys where data was captured before and after training, from staff who attended the training (Bull & Dale 2021, Chisholm et al. 2020, Hollis et al. 2021). Two studies undertook semi-structured interviews of staff involved in the implementation, delivery and evaluation of MECC training (Chisholm et al. 2019, Nelson et al. 2013). Two studies performed post-training online surveys of staff (Parchment et al. 2023) or nursing students (Mills et al. 2021) who attended MECC training. One study was a descriptive qualitative study which carried out focus group interviews of students who had attended training (Tuohy et al. 2021). Finally, three studies used several methods of data collection including training evaluations, surveys and interviews of staff who attended training (Lawrence et al. 2022, Pallin et al. 2022), and staff who both delivered and attended training (Dewhirst & Speller 2015).

**Table 1:**
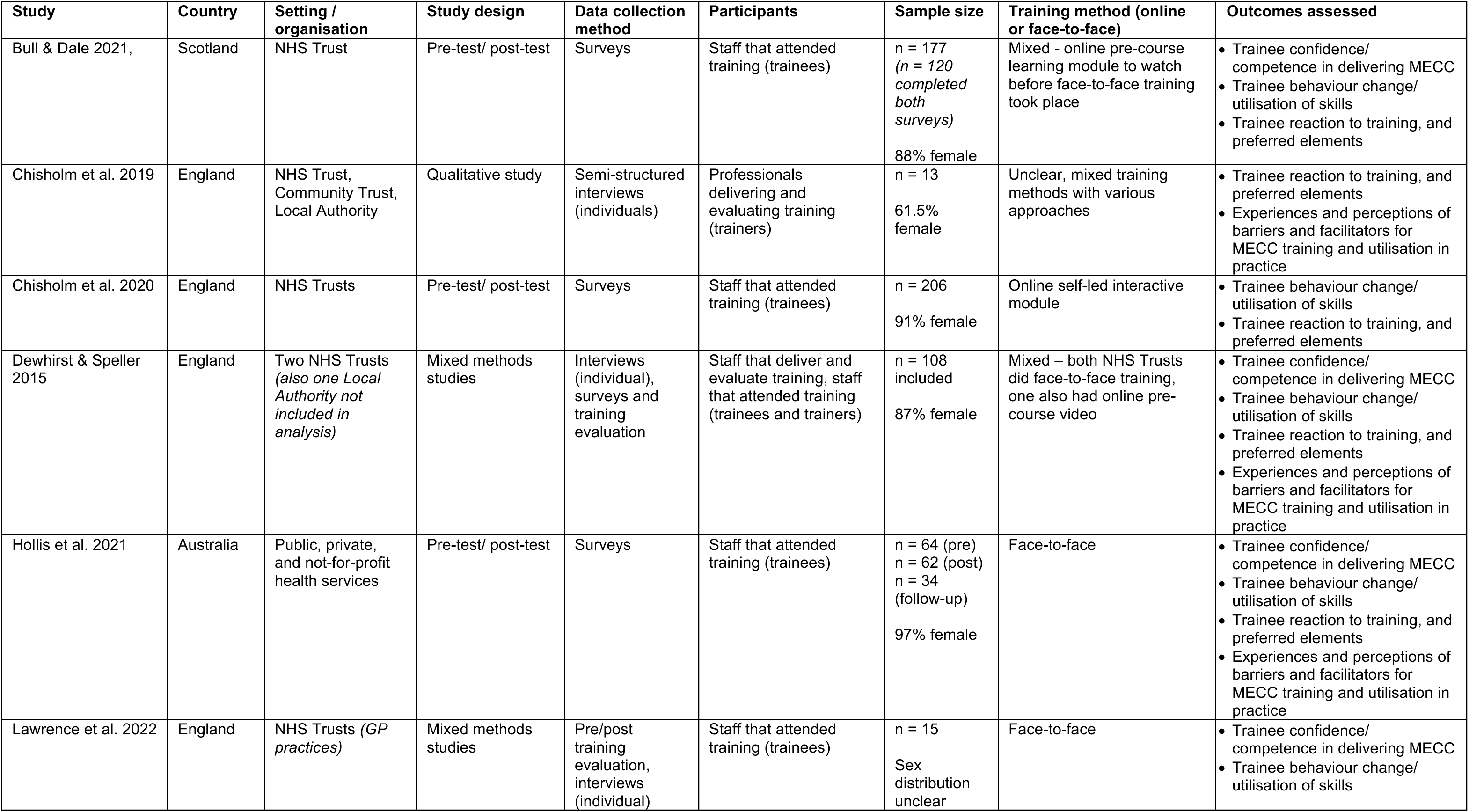
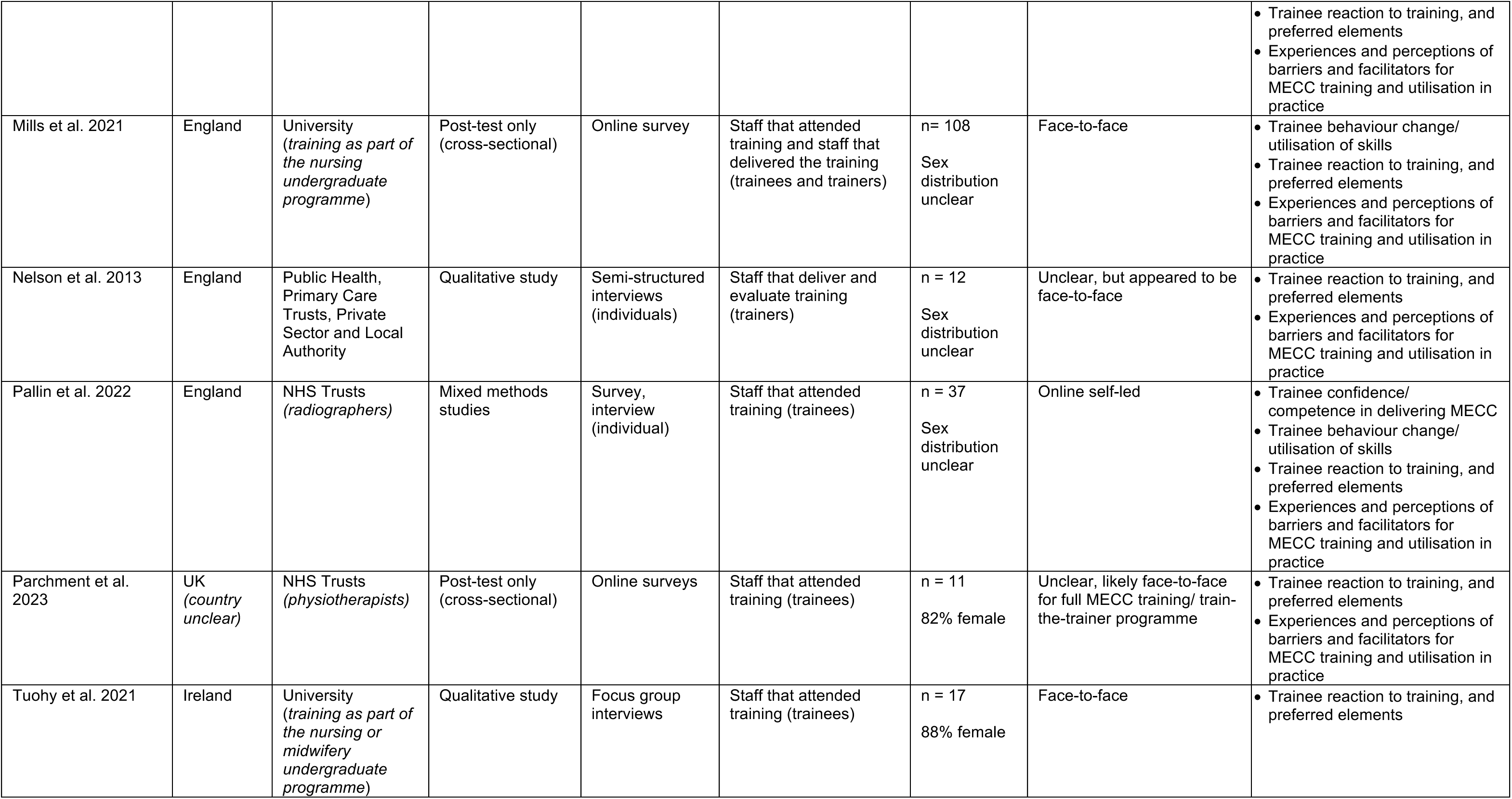
Study characteristics.

Throughout this review, staff or students who attended training will be referred to as ‘trainees’. This may include health-care professionals, practitioners, or other staff. People who delivered, implemented or evaluated training will be collectively referred to as ‘trainers’. This may include public health practitioners or other staff.

Dewhirst & Speller (2015) performed a pilot study of MECC implementation at three NHS Trusts, with pre-post surveys and semi-structured interviews. Lawrence et al. (2022) performed a longitudinal feasibility study with pre and post training evaluation surveys, observations, and interviews. Pallin et al. (2022) undertook a sequential mixed-method design, with participants completing two online courses and completing online surveys after each. The studies were conducted in England (n = 8), Scotland (n = 1), Ireland (n = 1) and Australia (n = 1). A detailed summary of the included studies and their findings are presented in Section 6, where studies reporting quantitative results are presented in Table 3, and studies reporting qualitative results in Table 4. Studies collecting both qualitative and quantitative data (mixed methods studies) are reported in both tables.

Two studies used online training only (Pallin et al. 2022, Chisholm et al. 2020), two studies did face-to-face training only (Lawrence et al. 2022, Hollis et al. 2021) and two included training as part of the nursing or midwifery undergraduate programme which was provided face-to-face (Mills et al. 2021, Tuohy et al. 2021). Three studies utilised mixed training methods, with two providing an online pre-course learning module to watch before face-to-face training took place. The online pre-course module was provided to everyone trained within the study reported by Bull & Dale (2021) and for those at one of the three NHS Trusts (Hampshire Hospitals Foundation Trust; HHFT) included by Dewhirst & Speller (2015). The training provided varied between the Trusts included by Chisholm et al. (2019). It was unclear whether training was completed online or face-to-face in the publications by Nelson et al. (2013) and Parchment et al. (2023)-there were indications that training was completed face-to-face, but as a variety of organisations were included and detail not provided on each individually, this may have varied. No studies were identified that provided training remotely via platforms such as Zoom or Microsoft Teams.

There was no comparative evidence reviewing different methods of MECC training identified. Therefore, this review is focused on the evidence around efficacy of different modes of MECC training, trainee response, and which elements of training were preferred by trainees or presented barriers to attendance or its perceived usefulness. There was also variability in outcome assessment and reporting between included studies, particularly with quantitative outputs, meaning direct comparison was not usually possible, and a meta-analysis could not be undertaken. There were issues with potential bias (which may distort the results) across the identified studies, with many using self-selecting samples and specific study populations, limiting the generalisability of results to the wider NHS Wales (see sections **Error! Reference source not found.** and 3.2, and Table 5 for further details). The approach used to review the methodological quality of included studies is outline in Section 5, which included the use of a tailored quality assessment tool.

#### 2.1.2 Effectiveness of MECC training

The type of outcome measures used for assessing the most effective method of delivering MECC training were divided into those that were considered by the stakeholders as essential (primary outcomes) and those that were considered important (secondary outcomes). The results of individual studies are provided in the tables presented in Section 6 (Table 3 and Table 4), and are summarised below.

##### Primary outcomes

###### Trainee confidence/ competence in delivering MECC

Five of the included studies reported quantitative results for change in trainee confidence and/or competence (Bull & Dale 2021, Dewhirst & Speller 2015, Hollis et al. 2021, Lawrence et al. 2022, Pallin et al. 2022).

Methods of measuring *trainee confidence* varied, but Bull & Dale (2021), Dewhirst & Speller (2015), and Hollis et al. (2021) all involved participants rating their own confidence in delivering MECC on a scale of 1-10 both prior to, and after training, and assessed the change in score. Bull & Dale (2021) asked participants to assess their confidence in relation to the following specific behaviour change techniques after completing the online pre-course module and face-to-face sessions: ‘information about health consequences’, ‘pros and cons’, ‘action planning’, ‘self-monitoring of behaviour’, and ‘prompts and cues’. Mean ratings of confidence for all behaviour change techniques increased following the training, to a statistically significant extent (p < 0.001). Dewhirst & Speller (2015) also reported an increase in the proportion of attendees feeling ‘very confident’ in raising the subject of healthy lifestyles with service users, with 24% of attendees initially reporting this, to 29% after participating in face-to-face training led by local leads who undertook ‘train the trainer’ courses (Dewhirst & Speller 2015). Confidence in having behaviour change conversations remained higher than it was before face-to-face training, despite a reduction from ratings reported immediately after training to 6-10 weeks follow-up (Hollis et al. 2021). It was also noted that trainees who give direct care to service users maintained higher confidence in supporting service users to make behaviour changes compared to those who did not (Hollis et al. 2021). Pallin et al. (2022) reviewed a variety of online courses and had trainees rate various statements on the impact of training on a scale of 1 to 5. Trainees agreed that training had given them confidence in having conversations about healthy eating (mean rating 4.0) and physical activity (mean rating 3.8). These results were not split out by type of training, so it was unclear whether confidence varied depending on whether the course was via videos, a website, or an online course.

Two studies reported qualitative results for change in *trainee confidence* (Chisholm et al. 2020, Nelson et al. 2013). Participants included by Chisholm et al. (2020) were asked to reflect on their practice, and they reported feeling that the online training enhanced their confidence to discuss lifestyle topics with service users. As a result of the training, most people reported that having MECC conversations was easier, although one individual felt that their confidence was undermined by the training as they found the content was too rigid. The MECC training delivered by some of the people interviewed in the study by Nelson et al. (2013) was aimed at people with no public health experience (e.g. working in the Local Authority), and thus a key focus was on building the confidence of trainees to have conversations with service users about their health and behaviours, as this was felt to be most beneficial.

*Trainee competence* in utilising behaviour change techniques, as measured by self-reported ratings out of 10, all increased to a statistically significant effect (p < 0.001) after face-to-face training with a pre-course online module, when compared to before (Bull & Dale 2021). Additionally, when Lawrence et al. (2022) undertook observations of participants in their use of the four areas of HCS (open discovery questions, reflection, listening and goal-setting), at clinic sessions 1-2 and 11-13 months after face-to-face HCS training, they found high levels of competence at both, indicating skills were maintained. When rated out of 5, participants in Pallin et al. (2022) who had undertaken training via online courses, noted an improvement in their knowledge on what to say to patients about healthy eating and physical activity, and also how to fit these conversations into the time available as part of their consultations. As for confidence outcomes, it was unclear whether competence reported by Pallin et al. (2022) varied depending on whether the course was videos, a website, or an online course.

###### Trainee behaviour change/ utilisation of skills

Six of the included studies (Bull & Dale 2021, Dewhirst & Speller 2015, Lawrence et al. 2022, Chisholm et al. 2020, Pallin et al. 2022, Mills et al. 2021) reported quantitative results for trainee behaviour change or utilisation of skills.

Participants in Bull & Dale (2021) rated their intention to use various behaviour change techniques (‘information about health consequences’, ‘pros and cons’, ‘action planning’, ‘self-monitoring of behaviour’, and ‘prompts and cues’) on a scale of 1 to 10 before and after completing the pre-course online module and face-to-face training. All intention ratings were quite high even before training, but most still increased by a statistically significant extent, with only ‘information about health consequences’ not reaching significance (p = 0.148). Chisholm et al. (2020) asked participants who had undertaken online training to rate their behavioural expectation. Participants did this by indicating how many service users they expected to have healthy behaviour conversations with, out of every 10 that they saw. The mean rating was 6.26, and the mode was 10, with 24% of users expected to have conversations with all service users, and the other 76% normally distributed around a mean and median of approximately 5 (i.e. expected to have conversations with about half of service users). Dewhirst & Speller (2015), Lawrence et al. (2022) and Hollis et al. (2021) counted the use of open discovery questions s before and after face-to-face training of varying durations. All three studies found an increase in use of open discovery questions after training, with two reporting statistically significant results (p < 0.001), and one not performing statistical analysis (Lawrence et al. 2022). Hollis et al. (2021) noted a statistically significant improvement in OQD use both immediately following face-to-face training and at 6-10 weeks follow up. The combined data for both NHS Trusts that included HCS elements also had a statistically significant improvement in use of open discovery questions following face-to-face training delivered by local leads (Dewhirst & Speller 2015). There was an increase in how often trainees said they actually raised the subject of healthy lifestyles with service users, from 39% attendees raising this at ‘most contacts’ or ‘every contact’ before training, to 56% afterwards (Dewhirst & Speller 2015), with a corresponding reduction of attendees ‘never’ raising healthy lifestyles. These differences were found not to be statistically significant. Ratings provided by Pallin et al. (2022) of participants who had undergone two online training courses focusing specifically on either healthy eating or physical activity, indicated they felt it was both appropriate to have discussions around these topics with their patients, and that they should do this as part of their professional role. Finally, Mills et al. (2021) reported that most students (66%) who received HCS training as part of their undergraduate course used the skills they had learned regularly or occasionally in practice. A further 25% of students reported that they hadn’t yet used the skills, but planned to when the opportunity arose, and the final 9% of students did not feel able to use the skills.

There were no qualitative results reported for this outcome.

##### Secondary outcomes

###### Trainee reaction to training, and preferred elements

Five included studies reported quantitative results for trainee reaction to training, and preferred elements (Bull & Dale 2021, Dewhirst & Speller 2015, Hollis et al. 2021, Mills et al. 2021, Pallin et al. 2022).

Participants rated various aspects of the training provided on a scale of 1 to 10 within studies reported by Bull & Dale (2021) and Dewhirst & Speller (2015), and a scale of 1 to 5 in the study reported by Hollis et al. (2021) and Pallin et al. (2022), where 1 was the lowest rating and 5 or 10 were the highest. The face-to-face training gained average ratings of 8.5 or higher for relevance, how interesting it was, and how well it met its objectives and the trainees learning needs in Bull & Dale (2021) (Table 3). Trainees rated the value of the face-to-face training undertaken in Dewhirst & Speller (2015) as an average of 9 out of 10. Trainees who attended face-to-face training in the study reported by Hollis et al. (2021) rated their satisfaction with the training as 4.9 out of 5. Mills et al. (2021) asked the students who received MECC training within their undergraduate degree to score the usefulness of the skills they had acquired in helping them start healthy conversations on a 3-point Likert scale of ‘very helpful’, ‘helpful’, or ‘not helpful’. They reported that 84% of students found the skills acquired in training ‘very helpful’ or ‘helpful’ in starting healthy conversations. Participants included by Bull & Dale (2021) rated the training as highly interesting, relevant, and that it met their learning needs. Participants in Pallin et al. (2022), some of which were from Macmillan and other charities, indicated that online training had made signposting to resources related to healthy eating and physical activity easier. Relevance of, and interest in the training were rated highly in the study by Bull & Dale (2021), with 92% of participants also rating the balance of course activity assigned to trainer talking time, and questions and discussion, as ‘about right’.

Nine studies reported qualitative results on trainees’ reaction to training and preferred elements (Bull & Dale 2021, Chisholm et al. 2019, Chisholm et al. 2020, Dewhirst & Speller 2015, Lawrence et al. 2022, Nelson et al. 2013, Pallin et al. 2022, Parchment et al. 2023, Tuohy et al. 2021). Following a thematic analysis, three themes and their subthemes were identified, these have been summarised in Figure 1 and expanded below.

**Figure 1:**
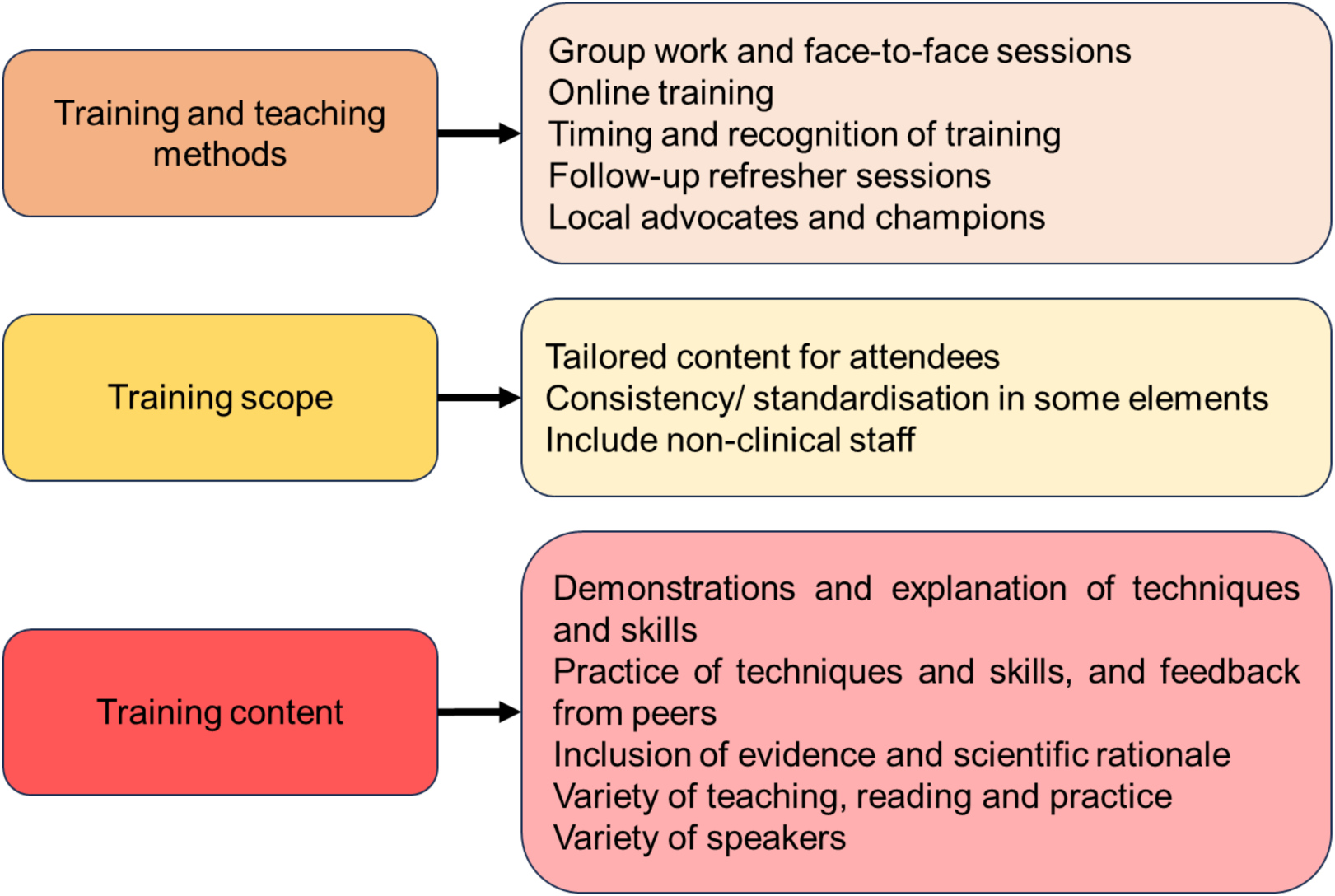
Themes and subthemes for training preferences.

The first main theme identified is ‘Training and teaching methods’. The included studies had a mixture of online and face-to-face sessions. Overall, the preference of attendees seemed to be face-to-face sessions, as this allowed for group work and improved focus as it reduced the number of distractions that may be experienced (Chisholm et al. 2020). However, there was recognition across several studies that there was a place for online training as this allowed more flexibility for trainees to undertake training at a time which was convenient for them. The timing and recognition of training was also frequently discussed. Several studies noted that making training mandatory could increase staff confidence in it, highlight its importance, and could increase support for training at management level (Pallin et al. 2022). There was a suggestion that including MECC training as part of staff induction, or even within undergraduate or other professional training would be useful to emphasise its importance (Parchment et al. 2023, Pallin et al. 2022, Nelson et al. 2013). Further to this, providing staff with protected continuing professional development (CPD) time and/ or accreditation following MECC training would also help emphasise that staff should be having these conversations with patients and improve managerial support (Nelson et al. 2013, Pallin et al. 2022).

The second theme was ‘Training scope’. There was agreement across the identified studies that it was useful to have consistency and standardisation in some elements of the training, but to ensure content can be specifically tailored for attendees. Although this would make development and delivery of training more complex, it does seem to mean attendees are more engaged, and get more from the training sessions. Finally, it was felt that it would be useful for both clinical and non-clinical staff to undergo training, as anybody who interacts with service users, including porters and receptionists, may be able to have MECC conversations with them (Nelson et al. 2013).

Finally, the third theme was ‘Training content’. Linked with the slight preference for face-to-face training, was a feeling that practice of techniques and skills, with peer feedback, following an initial demonstration and explanation, was a very useful element of training. Participants liked to have interactive and participatory training sessions, where they could be briefly informed about the background and rationale behind healthy behaviour conversation techniques such as behaviour change techniques and open discovery questions, before putting these into practice with their peers. These elements of interaction with peers, and practicing techniques, would not be possible with online self-led courses. Participants valued the inclusion of a brief summary of the evidence and scientific rationale behind MECC and the techniques being used, and felt this helped them feel more confident in the reasoning behind, and importance of, putting them into practice (Pallin et al. 2022, Tuohy et al. 2021). Participants also liked having a variety of speakers, with a mixture of teaching, reading and practice as part of the training-noting that there needs to be a variety of techniques used as everybody learns differently and the training should be developed to cater to as many of these requirements as possible (Tuohy et al. 2021).

###### Experiences and perceptions of barriers and facilitators for MECC training and utilisation in practice

Two included studies reported quantitative results around barriers and facilitators for MECC training and utilisation (Hollis et al. 2021, Mills et al. 2021). Prior to training, skills, beliefs about capabilities, intentions, goals, memory, attention and decision processes, and behavioural regulation were identified by Hollis et al. (2021) as potential barriers to having behaviour change conversations. These improved (i.e. were perceived to be less of a barrier) after the face-to-face training was provided, with three maintaining a statistically significant change (p < 0.01) from baseline to the last date of follow-up at around 6-10 weeks after the course: skills, belief about capabilities and goals (Table 3). Barriers in utilising HCS in practice were experienced by 29% of participants in Mills et al. (2021), and the most commonly reported were lack of time, lack of confidence, lack of knowledge of local services, lack of mentor knowledge or role modelling of HCS, and a feeling that HCS skills are not valued. Both studies also reported potential enablers to utilising MECC and/ or HCS in practice, noting that additional resources and information on local services available would be useful, as would regular training and refreshers (with mixed responses as to whether this should be online or face-to-face). There was some interest in having a ‘buddy’ to reflect with, and having training included in standard professional training academic modules, but less interest was shown in options such as including review of HCS in annual performance reviews, follow-up telephone meetings with a trainer, or connecting with other training participants via a social media group (Hollis et al. 2021).

Seven studies reported qualitative results on experiences of barriers and/or facilitators for MECC training and/or utilisation in practice (Chisholm et al. 2019, Dewhirst & Speller 2015, Lawrence et al. 2022, Mills et al. 2021, Nelson et al. 2013, Pallin et al. 2022, Parchment et al. 2023). Following a thematic analysis, four themes and their subthemes were identified, these have been summarised in Figure 2 and expanded below.

**Figure 2:**
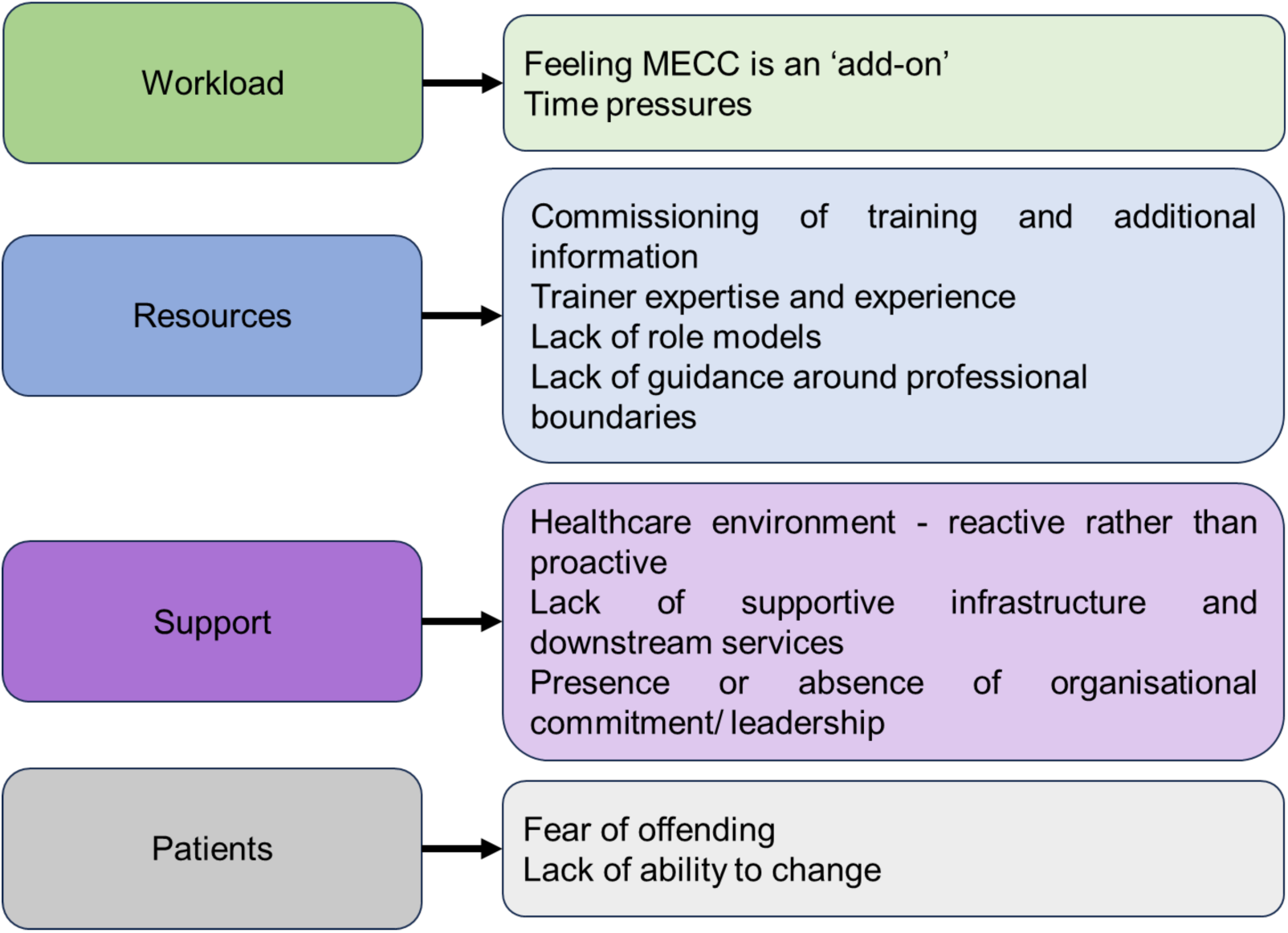
Themes and subthemes for barriers and facilitators of MECC training and utilisation.

The first identified theme was the ‘workload’ of trainees in the included studies. The most regularly occurring barrier to utilising MECC was the time pressure of their workload, and feeling MECC was an ‘add on’ to this. Trainees did not feel they had sufficient time or support to have MECC-based conversations with service users. Potential enablers linked to this have been summarised within the next themes.

The second theme identified was ‘Resources’. Participants in the included studies often said that the commissioning of training and provision of additional information would help highlight the importance of MECC both locally and more widely. In doing this, more people would feel supported in, and able to, have MECC conversations with service users. Related to this, was the feeling that having trainers with adequate expertise and experience was also important: ‘train the trainer’ models only worked if those who cascaded the training locally felt competent and confident in MECC and the various techniques being taught within the training modules (Nelson et al. 2013, Parchment et al. 2023). If trainers were not deemed to be experts, it was felt to undermine the training and importance of the techniques being introduced (Dewhirst & Speller 2015). Additionally, the presence of role models was seen to be an enabler, with the lack of local champions or role models seen as a barrier to implementing MECC in practice. An absence of guidance around professional boundaries was also seen to be a barrier, and the provision of such guidance was felt to increase people’s confidence in MECC and help them feel more supported in their utilisation of it in practice.

Thirdly was the theme of ‘Support’, primarily linking with the presence or absence of supportive infrastructure and organisational commitment for MECC training and utilisation. Many participants said a big barrier to using MECC was a lack of knowledge, or presence, of downstream services to refer service users to following healthy behaviour conversations (Mills et al. 2021). It was also mentioned that the healthcare service as a whole is seen by many trainees to be reactive rather than proactive, and a perspective shift would be needed to enable more to feel confident and comfortable having MECC-based conversations with service users (Chisholm et al. 2019). Finally, the presence or absence of organisational commitment to MECC training and utilisation, and leadership within organisations, was seen to have a big role in whether people felt able and supported to have conversations with service users. People working in organisations without support from management felt unable to have healthy behaviour conversations, and having a supportive management team made a big difference to how people felt about putting their training into practice.

The final theme identified was the patients or service users themselves. Trainee perceptions of the service users had a big impact on whether they felt comfortable having healthy behaviour conversations with them. There was a recurring theme of trainees being afraid to cause offence to service users, and, less commonly, a concern that the service users lacked the ability to change due to personal circumstances or attitudes (Lawrence et al. 2022). Having a fear of offending service users, or not feeling that they would be able to make changes even if they wanted to, prevented trainees feeling comfortable in having MECC conversations and caused a large barrier to utilisation of MECC. However, there was a feeling that some of the techniques introduced in MECC or HCS training (both face-to-face and online) such as behaviour change techniques and open discovery questions could help overcome this fear, and with practice, participants felt they would be more confident in using these in their day-to-day interactions and have less fear of offending patients.

###### Rates of signposting/ referral and uptake of these services

None of the included studies included data for this outcome.

#### 2.1.3 Quality of included studies

A detailed summary of the assessment of the quality of individual studies is provided in Section 6.3 (Table 5). There were several studies in which concerns about study representativeness were flagged. Most studies identified people by invitation with eventual participants being self-selecting and both willing and supported to do the training and put the learned techniques into practice. In addition to this, although some studies included participants with several different job roles, most were in specific populations such as General Practitioners (GPs) and radiologists. Finally, many of the included studies had a high proportion of female participants. As such, the generalisability from the results reported is limited, as only some job roles were covered, and within the NHS more widely there may be variable enthusiasm and willingness to implement MECC in practice. Further to this, the included departments had local support for MECC or HCS training, which again may limit generalisability as it may be that not all NHS Trusts and departments are supportive of the approach/ feel they have capacity to release staff to undertake the training.

Another consistently flagged issue was the potential for non-response bias. As most information was collected via interviews or surveys, response rates varied, and some studies had higher risk of non-response bias than others. This could particularly be the case where surveys were collected a long time after training, and one study (Hollis et al. 2021) did not report qualitative data due to low uptake.

Overall, the combining of qualitative and quantitative data within the studies was acceptable, if done to variable degree, and the data complemented each other. There were some consistent themes and results across the studies, so despite the issues with generalisability and some conflicting outcomes around training preferences, the evidence identified may still be useful in the commissioning, planning and delivery of MECC training.

#### 2.1.4 Bottom line results for MECC training

The available evidence did not allow for comparison of different methods of MECC training. Therefore, this review focused on the evidence around efficacy of different modes of MECC training, response to training, and trainees preferred elements.

Overall, the identified evidence suggests trainee confidence and competence increased following MECC training, regardless of whether this was online or face-to-face. There was less evidence regarding behaviour change and utilisation of skills in practice, however, from the evidence that was available, it was suggested that MECC training has a positive effect and these increase following training. There was less consistent evidence on training preferences; some participants indicated a preference for online training as it was more flexible, but most preferred in-person training as it allowed practice of the skills and discussion with peers. There was an indication that additional support such as allowing CPD time for training or introducing an accreditation, and greater support from management and the host organisation, would be useful in emphasising the importance of MECC and increase uptake of MECC training and utilisation in practice.

## 3. DISCUSSION

### 3.1 Summary of the findings

The aim of this review was to assess which elements or forms of training for MECC, or comparable interventions, are most effective and which are preferred by those who would implement MECC in practice. The evidence indicates that training can be effective in improving trainee confidence and competence and can increase the number and quality of MECC conversations they have, or are likely to have, in practice. However, several barriers are still present in the workplace and these need to be overcome to result in culture change and widespread implementation.

#### Trainee confidence and competence in delivering MECC

There was consistent evidence across the identified studies that self-reported confidence and competence in having MECC-based discussions with service users improved following both face-to-face and online training. There were statistically significant improvements seen consistently for all face-to-face courses, regardless of duration and for both specialist MECC trainers and local leads who had undergone train-the-trainer courses. However, there was no statistical analysis undertaken for the online courses identified, so it was not possible to see whether the improvements noted were statistically significant. There was also some evidence that improvements in competence and confidence in delivering MECC of trainees were maintained for at least several weeks, but no evidence on the long-term impact. No confidence or competence results were reported for studies which included MECC or HCS in standard professional training.

#### Trainee behaviour change and utilisation of skills

There was an indication that training resulted in an increase in both trainee expectation of having healthy behaviour change discussions with service users, and actual conversations undertaken. However, this was variable, with some trainees anticipating having conversations with most service users, but the majority expecting to have conversations with around half of the service users they met with. There was some evidence indicating that the use of open discovery questions increased after training, but no evidence on the long-term impact of training on trainee behaviour change and skill utilisation, with the maximum follow-up of the included studies being 11-13 months after training.

#### Trainee reaction to training and preferred elements

There was consistent evidence that training was seen to be useful and relevant by trainees, but these were mostly self-selecting samples who already had an interest in MECC which limits the generalisability of this result. There was mixed evidence on whether online or face-to-face training was preferred, but a strong indication across all included studies that trainees liked practical and interactive training, with group work and time allowed for practice. Additional to this, there was support for some standardisation of some elements of training but with scope for it to be tailored to the needs of the attendees, and that regular refresher training would be beneficial. There was a reasonable amount of evidence suggesting that trainees felt training should be mandatory, and some suggestion that it may be beneficial to include it as part of professional qualifications or induction for both clinical and non-clinical staff, as they felt that this would increase staff confidence in the training as well as improve managerial support for it.

#### Experiences and perceptions of barriers and facilitators for MECC training and utilisation in practice

There was a strongly emerging theme, consistently seen across studies, that trainees felt that they did not have enough time to have MECC conversations with service users. There was also an indication that a lack of knowledge about local services and infrastructure was an additional barrier to having healthy behaviour conversations with service users. There was some evidence that improving awareness of local downstream services and having ‘MECC champions’ to support and educate colleagues would facilitate increased support for MECC, and a strong suggestion that an increase in managerial support would help trainees to feel encouraged and able to have healthy behaviour conversations.

#### Rates of signposting/ referral and uptake of these services

There was no evidence identified for this outcome.

### 3.2 Strengths and limitations of the available evidence

Participants in many of the included studies were self-selected, whereby they voluntarily attended training or completed interviews or surveys. This could result in selection bias which could affect the validity and generalisability of the study findings. It may also result in an underestimation of the effect of the intervention, as those participating in the studies already had high levels of intention to use MECC, and thus any possible increases because of training could be reduced, as there was less improvement to be had. Conversely it could result in an increase of effect, as the participants would attend training with the intention of getting as much out of it as possible.

There was some variability in how outcomes were assessed across the literature, particularly the quantitative measures of confidence and competence, meaning that a meta-analysis (combining the results from multiple similar studies) was not possible. Additional to this, the timing of survey provision also varied, however the general themes and outcomes reported appeared similar across studies. There was also variation in what information about the training undertaken was reported, with a lack of clarity in some instances. Where multiple courses or training durations were included, there was not often a breakdown of differences in outcomes by course which meant it was not possible to assess exactly which were more effective, or preferred, compared to others.

Many of the pre-post studies collected data immediately before and after training. Feelings of confidence and intention are likely to be highest immediately after training, and this can often reduce once people return to work and face barriers to implementation (Bull & Dale 2021). No evidence was identified on the longer-term implications and effects of MECC training, other than the indication that there did seem to be a desire for refresher training, so the long-term effect is unclear.

Finally, none of the identified evidence assessed whether different methods of MECC training influenced the rates of referrals of service users to, and uptake of, specialist services. There is a lack of comparative evidence reviewing multiple MECC training methods and levels, and uncertainty around the impact of improving the MECC training on service user experience and outcomes.

### 3.3 Strengths and limitations of this rapid review

A strength of this review is that it included a systematic and comprehensive search of the available literature, including grey literature. Identified studies reviewed the effectiveness of training, preferred elements of training, and its impact on trainee use of MECC in practice. However, no evidence was identified which directly compared varying methods of MECC training, so it was not possible to assess which training methods are most effective via comparative evidence.

Ten of the eleven included studies were UK based, and therefore relevant to the Welsh setting, but none appeared to be set in, or include participants from Wales specifically. Paediatric settings were excluded, therefore any studies looking at MECC training and utilisation in the family setting were excluded and the results identified here may not be directly applicable. Additional to this, there were variable frameworks and ‘levels’ of training defined across the studies, sometimes with variable elements included in each, meaning it was not possible to directly compare the interventions.

Quantitative results were reported narratively, and qualitative data was reported via a thematic synthesis, where feasible. Generally, the results were consistent and complementary across both data types, with no major contradictions or divergence between them. Whilst the outcomes of interest in this review were reported by several included studies, the study methods and methods used or measuring the outcomes varied, meaning that a meta-analysis was not possible, and the review was limited to a narrative synthesis.

We used a published tool (the Mixed Methods Appraisal Tool (MMAT); Hong et al. 2018) to assess risk of bias in all included studies. This was developed and intended as a tool for assessing studies with a mixed-methods design; not all our included studies strictly met this definition. However, we judged it inappropriate to use a mixture of critical appraisal tools for different study types as this would hinder drawing overall conclusions about common risks of bias. After piloting the MMAT tool, we judged that it was adaptable enough to be used for all the study designs we included.

### 3.4 Implications for policy and practice

This review highlights that MECC training can be beneficial in improving trainee confidence and competence in using MECC in practice, but highlights that there are consistent barriers which are experienced and can prevent techniques learned being implemented. Several potential methods of overcoming these barriers have been identified and summarised, but the practicalities of implementing these remain unclear, and many do require a widespread culture change and significant opinion shifts.

There was no evidence identified as to whether changes to MECC training have an impact on service user behaviour change and eventual outcomes. This lack of evidence may mean that staff who like a good evidence base for changes in their behaviours, or implementation of new practices, are less willing to attend MECC training and implement it in practice. This is particularly relevant as some changes, including a shift to face-to-face training, may have an impact on its cost, both in regard to providing the training and allowing staff time to attend.

There were some indications that generalised online training was of use in some circumstances and was more convenient, but face-to-face training was preferred and found to be both more useful and allowed for some tailoring to increase its applicability to the trainees. Current Level 1 training in Wales is online, generalised, and consistent across the nation. This provides an opportunity to provide background information and evidence for MECC, as an introductory course. Level 2 training in Wales varies by health board, which allows for it to be tailored to local needs, and be face-to-face, potentially improving its effectiveness and support at a local level. Having a general guideline and template for level 2 training would ensure consistency across Wales whilst allowing for tweaks to improve relevance to the local setting. Linking the level 2 training back to the initial online level 1 course, and building upon lessons learnt whilst allowing for group work and practice may be an effective method of providing MECC training across Wales.

### 3.5 Implications for future research

There needs to be further research into how MECC use and effectiveness in practice changes following different types of training. Research into whether MECC training impacts upon the number of referral rates of service users to secondary services, their uptake of these referrals, and whether this varies for different population groups or intended target of the MECC conversation (e.g. weight, alcohol use, vaccines), would be of use. Additional to this, research into whether there is any long-term impact of various types of MECC training on service user behaviour change and eventual outcomes would likely widen support for it amongst staff. Finally, understanding service user acceptability of MECC would help overcome trainees concerns around upsetting patients, as this was a key barrier in the implementation of MECC which was identified within this review.

Research specifically into the MECC training being implemented across Wales, how staff interact with and use this, and what they find useful, relevant, and usable would also be worthwhile. Although some of the evidence identified in this review could be applicable to the Welsh context, as the research was undertaken in the UK and was based on the same theories and practices as MECC, it is important to understand what works and does not work for the users who are trained within a certain system and setting. Although this review summarised some elements of training which may be preferred by trainees, there was no comparative evidence, so undertaking comparative research may be useful. This could help improve what is offered to staff and, in turn, could improve uptake and enthusiasm for MECC. Inclusion of staff who are not already enthusiastic, and do not necessarily have managerial support for, utilisation of MECC in practice would be useful and potentially more reflective of the breadth of experiences in NHS Wales overall.

This review summarised some of the barriers and enablers for MECC training and utilisation, and identified some suggestions on how the barriers may be overcome, but did not assess the effectiveness of these. Further research into methods to overcome barriers, enhance the enablers of MECC training and utilisation, particularly around encouraging organisational and managerial support, would be useful.

### 3.6 Economic considerations^*^

The Public Health England MECC evaluation guidance 2020 recommends producing a business case for MECC programmes that includes the costs of delivery and considers the value for money of its implementation. The guidance proposes a Social Return on Investment (SROI) approach could be taken to evaluate a wider set of social, economic and environmental outcomes of implementing a MECC programme {Public Health England, 2020 #22}.

Most policy documents discussing the cost-effectiveness of MECC programmes cite the National Institute for Health and Care Excellence (NICE) public health guidance on Behaviour change: individual approaches [PH49] published in 2014. This guidance does not directly refer to MECC programmes, but it does report that brief behaviour change or signposting interventions addressing smoking, diet, physical activity, alcohol, sexual health and multiple health targets fall well below their cost-effectiveness threshold of £20,000 to £30,000 per Quality Adjusted Life Year gained {National Institute for Health and Care Excellence, 2014 #23}.

Three evidence reviews (a review of current NICE guidance and recommendations, a review of economic evaluations of behaviour change interventions, and a review on their effectiveness) informed the [PH49] guidance. The reviews focussed on interventions addressing six behaviours: smoking, diet, physical activity, alcohol, sexual health and multiple health targets. Interventions identified by the reviews fell well below the NICE threshold for cost-per Quality Adjusted Life Year (QALY), [£20,000 to £30,000 per QALY gained]. Further, the reviews state that interventions that targeted the general population exhibited better cost–utility results and were more likely to be cost effective than those aimed directly at vulnerable populations. However, the reviewers suggest the findings are to be interpreted with caution given the heterogeneity of identified economic analyses, lack of complete information and potential reporting biases {National Institute for Health and Care Excellence, 2014 #23}.

## Data Availability

All data produced in the present study are available upon reasonable request to the authors

## Abbreviation

BCT: Behaviour Change Techniques
CI: Confidence Interval
CPD: Continuing Professional Development
CTMUHB: Cwm Taf Morgannwg University Health Board
GP: General Practitioner
HCS: Healthy Conversation Skills
HHFT: Hampshire Hospitals NHS Foundation Trust
MECC: Make Every Contact Count
MMAT: Mixed Methods Appraisal Tool
N/A: Not Applicable
NCD: Non-Communicable Disease
NHS: National Health Service
ODQs: Open Discovery Questions
PCC: Portsmouth City Council
PDR: Personal Development Review
PLBC: Prevention and Lifestyle Behaviour Change: Competence Framework
SD: Standard Deviation
SHFT: Southern Health NHS Foundation Trust
SMARTER: Specific, Measured, Action oriented, Realistic, Timed, Evaluated and Reviewed
TDF: Theoretical Domains Framework
TeNT PEGS: T = Taking down barriers; EN = Changing the ENvironment; Th = Addressing Thoughts and emotions; P = Perform and practice; E = Empowering people to change; G = Achieving Goals; S = Social support

## 5. RAPID REVIEW METHODS

### 5.1 Eligibility criteria

**Table 2:** Eligibility Criteria.

|  | **Inclusion criteria** | **Exclusion criteria** |
| --- | --- | --- |
| **Setting** | Healthcare (could include hospitals, GP surgeries, pharmacies, other medical appointments such as pre-surgical visits)<br>Charities/ voluntary/ not-for-profit organisations<br>Other non-healthcare settings<br>Opportunistic one-to-one conversations with adults ($\geq 18$) | Conversations in paediatric settings ($<18$)<br>Group settings |
| **Perspective** | People involved in the commissioning or training of MECC, and those utilising MECC conversations in practice | Service user |
| **Intervention / exposure** | Varying types of MECC training (level 1 alone, or 1 and 2)<br>Training in other similar interventions which utilise key elements of MECC as per the Welsh MECC definition* | Training in non-MECC interventions (i.e. any that are not opportunistic brief behaviour change conversations, such as motivational interviewing, goal setting or reductionist approaches)<br>Evaluations of the effectiveness of MECC alone (i.e. without assessing training elements)<br>Planned conversations/ any interventions including screening questionnaires |
| **Comparison** | Varying types of MECC training (level 1 alone, or 1 and 2)<br>If limited data comparing different methods of MECC training are available, studies where the comparator is no intervention (or a non-comparative study) may be included |  |
| **Evaluation (outcome measures)** | *Primary outcomes:*<br>Trainee confidence and/or competence in delivering MECC<br>Trainee behaviour change and utilisation or application of MECC training in practice<br>*Secondary outcomes:*<br>Trainee reaction to training, and preferred elements |  |
|  | Experiences and perceptions of trainee's barriers and facilitators for MECC training and utilisation in practice<br>Rates of signposting and/or referral of service users to secondary services, and uptake of these/ impact on service user behaviour change |  |
| **Study design** | Systematic reviews/ scoping reviews<br>Comparative observational studies<br>Qualitative studies<br>Before/ after studies<br>Service evaluations (*UK based only*) |  |
| **Countries** | The findings from studies that align most closely to the Welsh MECC definition will be included, and generalisability to the Welsh context evaluated as part of the review with any that are not at all comparable being excluded. |  |
| **Language of publication** | English |  |
| **Publication date** | No limits |  |
| **Other factors** | None |  |

\*MECC: opportunistic brief intervention conversation. "Brief Intervention" defined as a short, structured conversation that offers opportunistic advice, discussion, or negotiation that aims to strengthen a person's commitment for behaviour change (NICE, 2006). It is a person centred – holistic approach, and is not Motivational Interviewing, goal setting or a reductionist approach.

### 5.2 Literature search

Medline and Embase via Ovid, CINAHL and Scopus/ Web of Science were searched, Overton was also searched using an amended strategy (limiting terms to ‘making every contact count’ only). Searches were performed between 26^th^ and 27^th^ June 2024, with Medline and Embase results going up to, and including, 25^th^ June 2024. See Section 8 (Appendix: Search strategies) for the search strategies and results.

### 5.3 Study selection process

Two reviewers dual-screened 200 of the titles and abstracts independently. Disagreements were settled by discussion and consensus. Agreement reached the 80% agreement threshold, so the remaining titles and abstracts were screened by the primary reviewer alone. Following this, 20 percent of all full texts were dual screened. As the agreement threshold (80%) was reached, the remaining records were screened by the primary reviewer alone. During independent screening, the primary reviewer consulted with the secondary reviewer in the case of any uncertainties.

### 5.4 Data extraction

The following data were extracted where available:

- Study information (author, year, country, study type)
- Population characteristics (including sample size, setting)
- Intervention characteristics
- Outcomes, outcome measures and data collection methods
- Findings including change in ratings using Likert scales, qualitative summaries, change in practice

Data were extracted by a single reviewer and quality assured by a second reviewer.

### 5.5 Study design classification

There were a wide range of study types included within this review, including qualitative, quantitative and mixed methods studies.

### 5.6 Quality appraisal

Due to the varying nature of the study design of included studies, the Mixed Methods Appraisal Tool (MMAT) (Hong et al. 2018) was used to assess their quality. This ensured one consistent methodology was used across the included studies, to ensure the outcomes and considerations were comparable and could be summarised clearly, despite the variable study types included.

### 5.7 Synthesis

Following extraction, quantitative data was narratively synthesised and analysed. It was not possible to conduct a meta-analysis due to the heterogeneity of study outcomes and outcome measures.

A thematic synthesis approach was undertaken for the qualitative data identified within this review. This was undertaken by identifying and summarising key words and topics in the included studies and grouping these into overarching themes and related subthemes within Microsoft Excel. A separate thematic synthesis was planned for each outcome measure (outlined in Table 2), but where there were only two or less studies reporting on the specific outcome a narrative synthesis was conducted instead.

The principles of the Kirkpatrick training evaluation model {Kirkpatrick Partners, 2024 #24} were used when agreeing upon the outcome measures and synthesising results in this review. There are four levels to this model;

1. Reaction: how favourable, engaging and relevant the trainees found the training.
2. Learning: the degree to which training enabled trainees to acquire the intended knowledge, skills, attitude, confidence and commitment.
3. Behaviour: the degree to which what was learned in training is applied in practice.
4. Results: the degree to which organisational outcomes occur following training.

For MECC, level four would be assessed via the number of signposting and referrals of service users that take place, and the uptake of these as well as actual behaviour change from service users.

## 6. EVIDENCE

### 6.1 Search results and study selection

**Figure 3:**
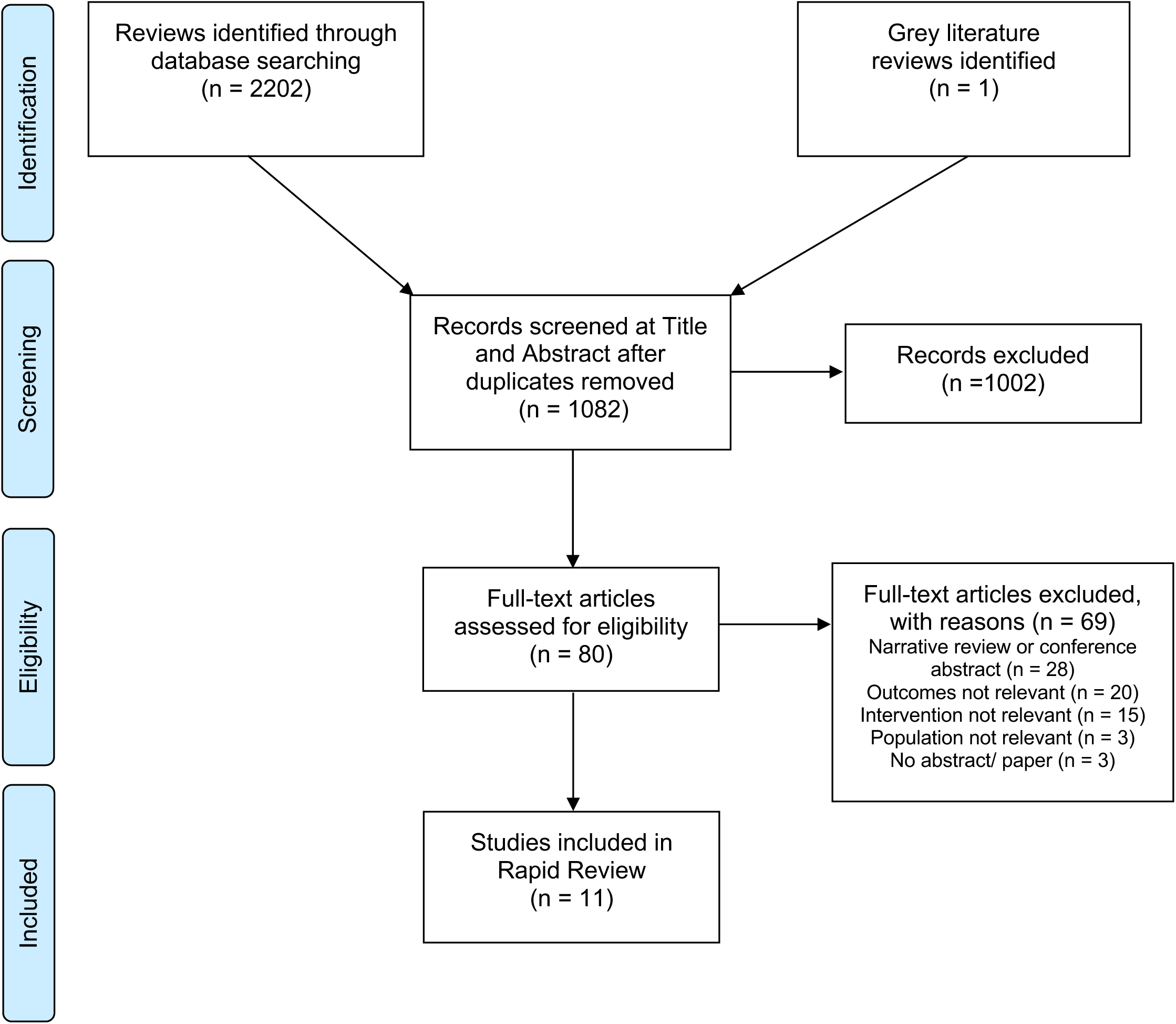
PRISMA flow diagram.

### 6.2 Data extraction

All 11 included studies are summarised between Table 3 and Table 4.

**Table 3:**
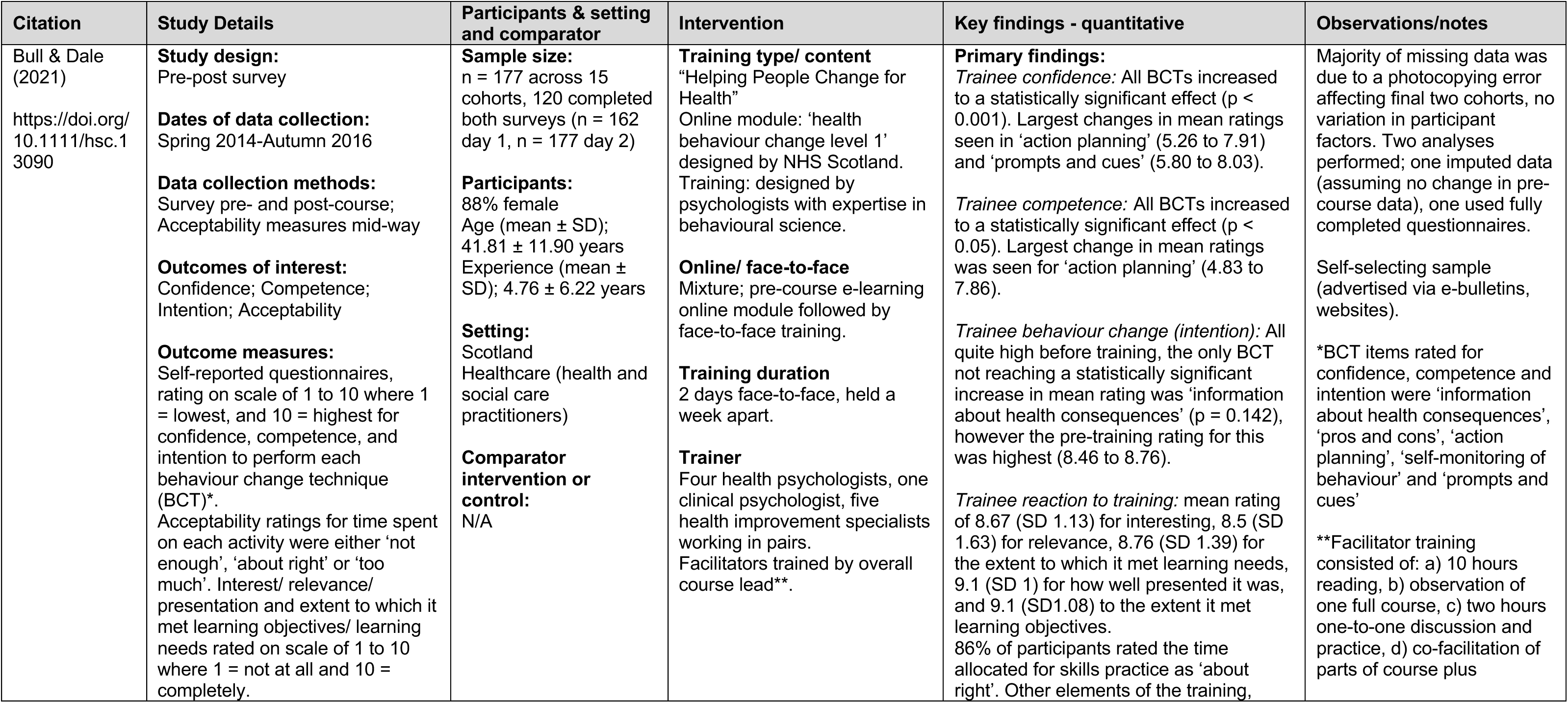
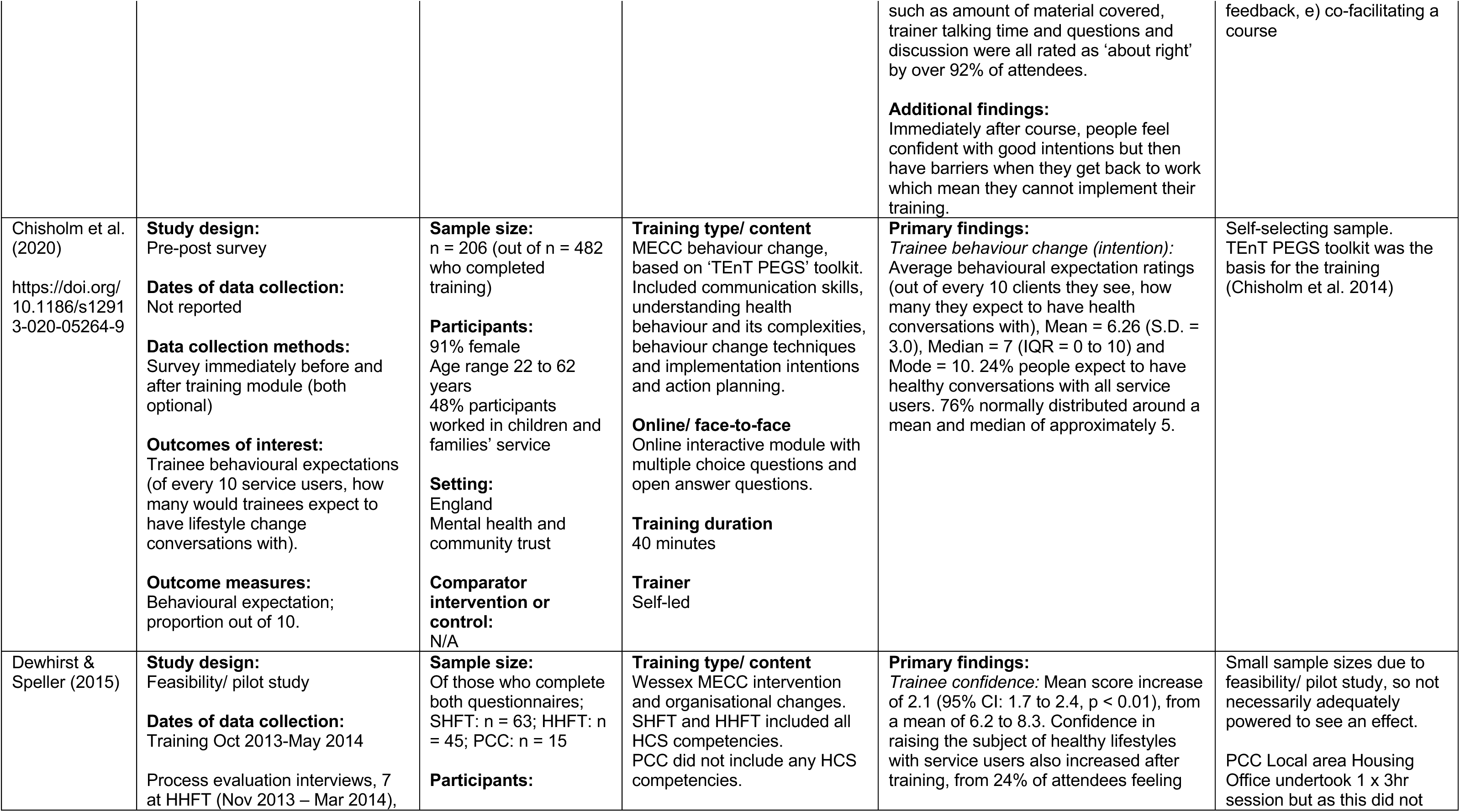
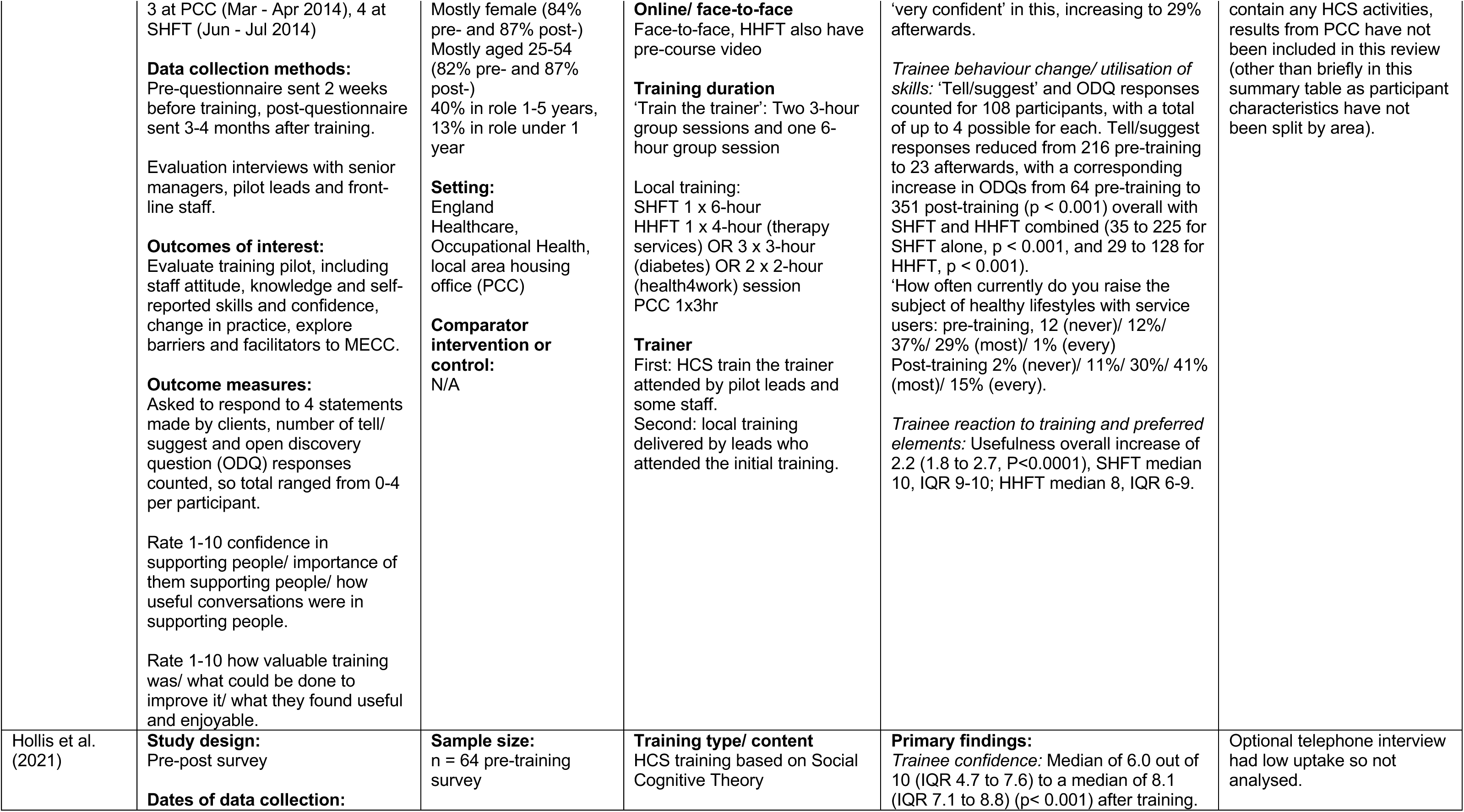
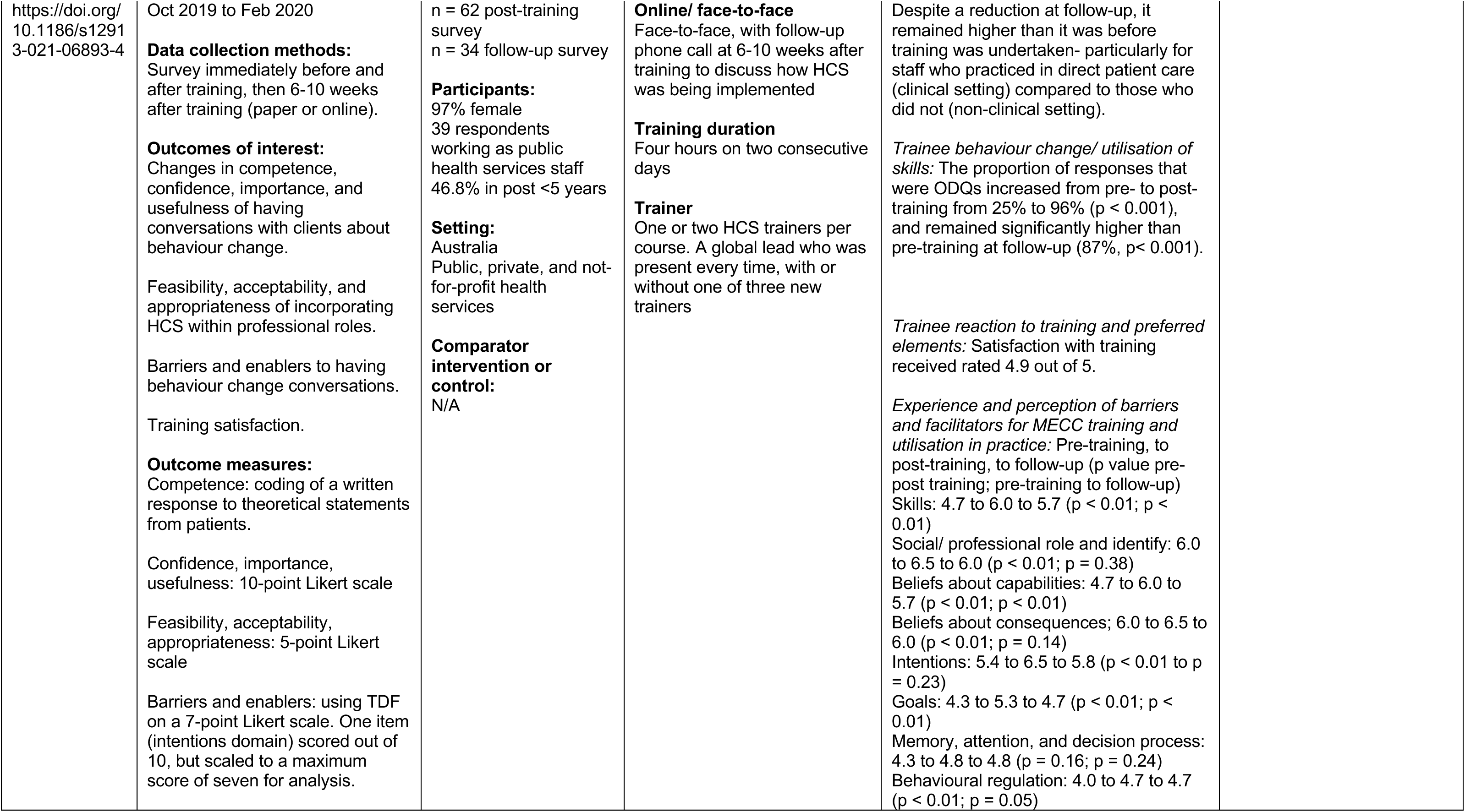
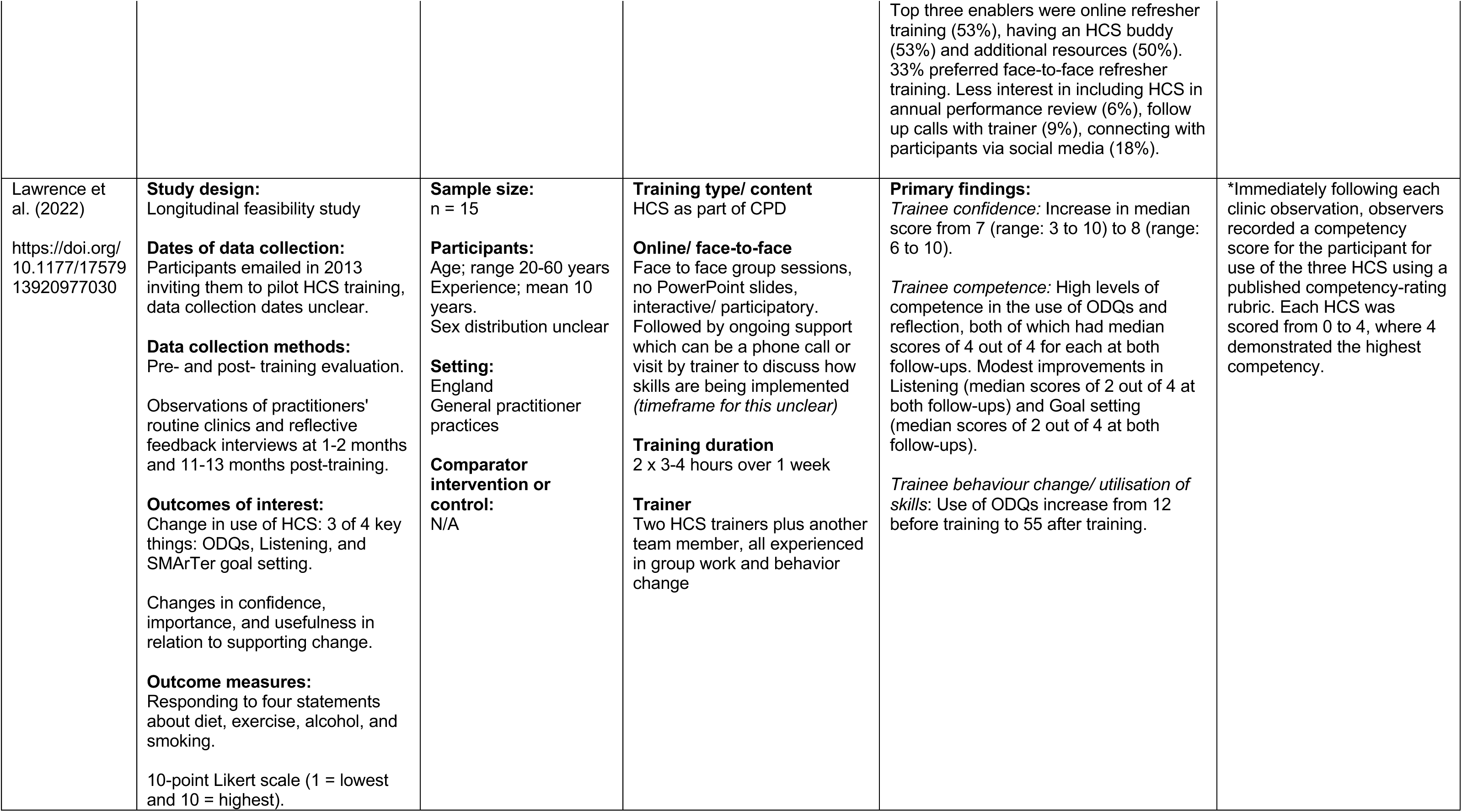
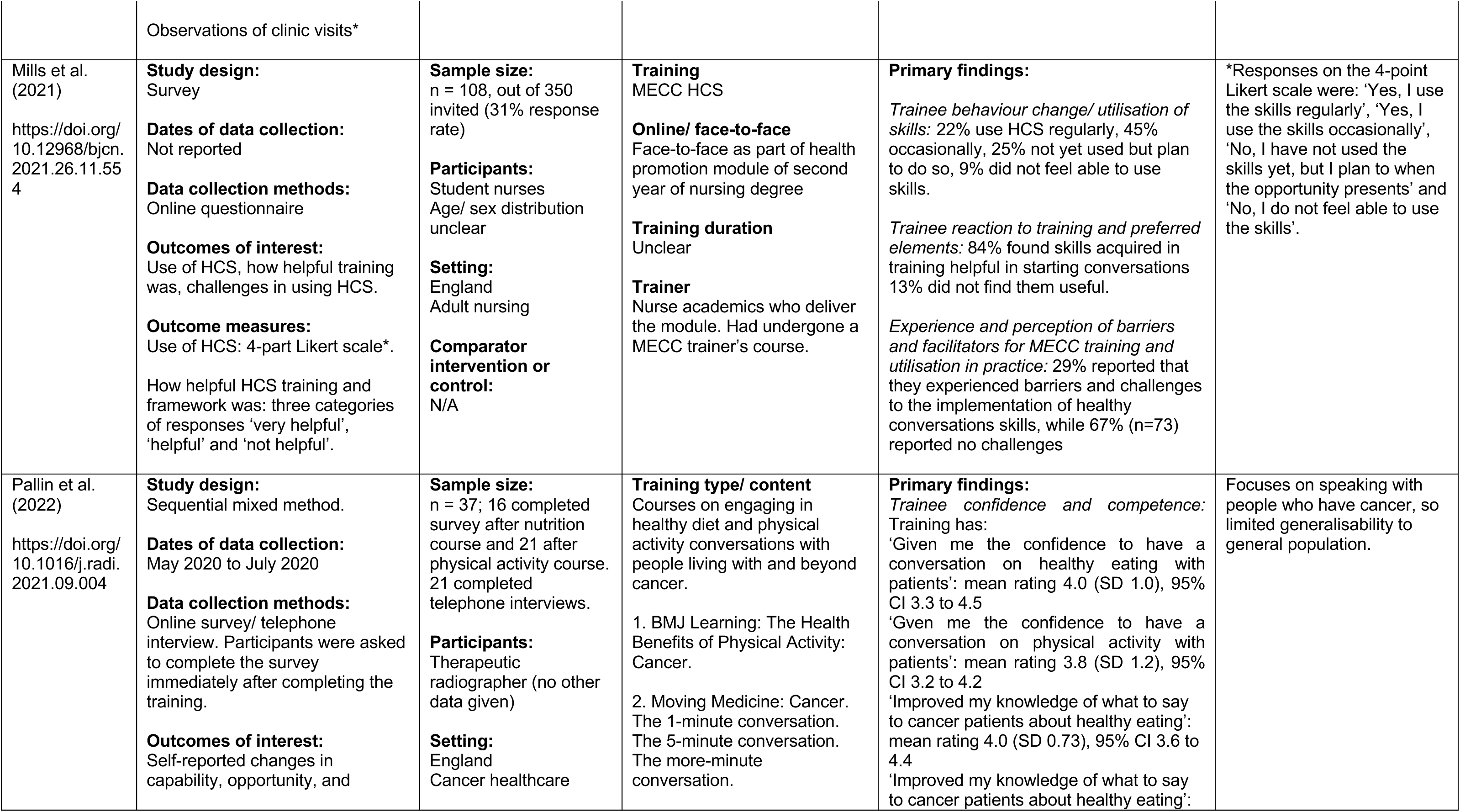
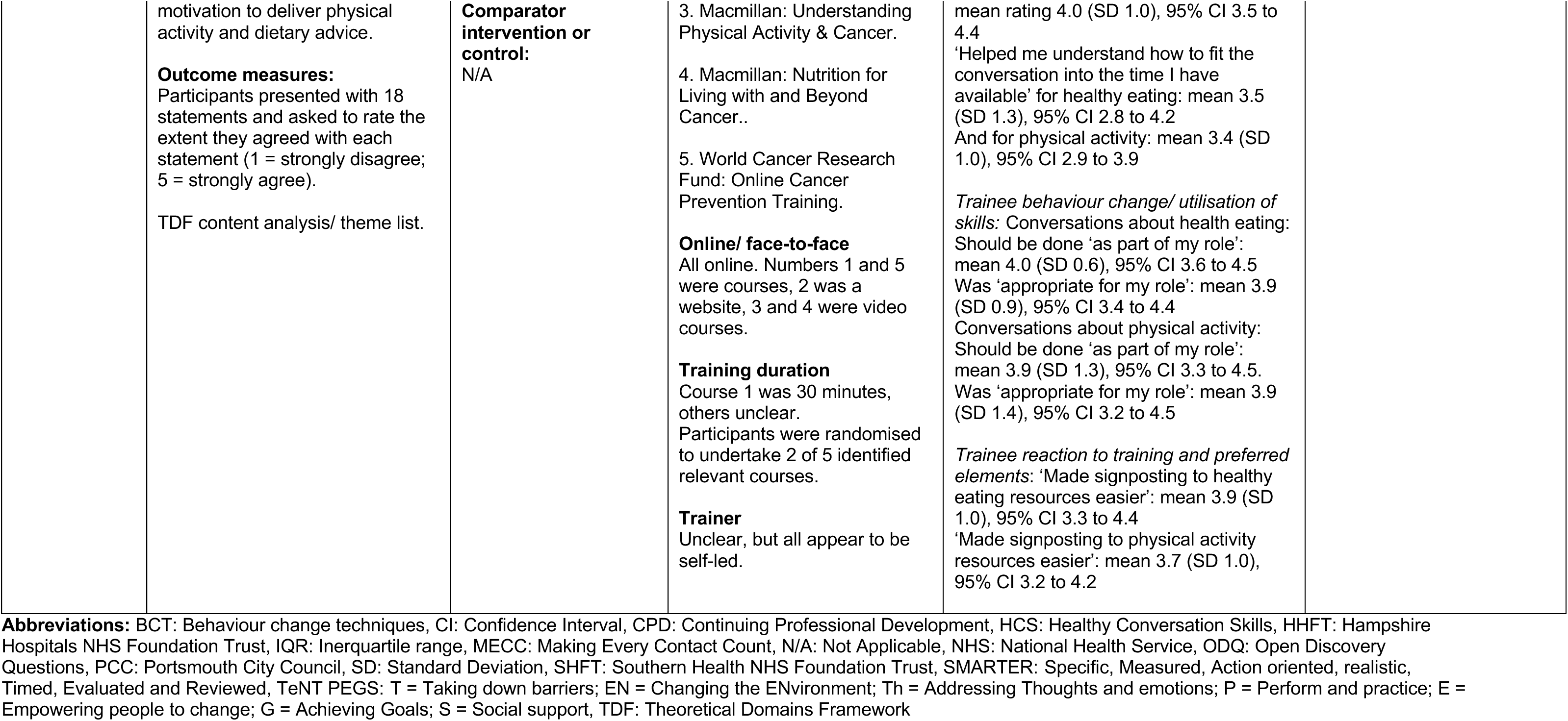
Summary of included studies and quantitative results. NB: only mixed methods or quantitative only studies are included in this table, qualitative studies and results are summarised in Table 4.

**Table 4:** Summary of included studies and qualitative results. NB: Full study details are noted in Table 3 for mixed methods studies, and only a summary of key details is included below-this is indicated in the ‘Observations/ notes’ column where it is the case. Full study details for qualitative studies are included below and not in Table 3.

| Citation | Study Details | Participants, setting and comparator | Intervention | Key findings - qualitative | Observations/notes |
| --- | --- | --- | --- | --- | --- |
| Bull & Dale (2021)<br>https://doi.org/10.1111/hsc.13090 | **Study design:**<br>Pre-post survey<br>**Dates of data collection:**<br>Spring 2014-Autumn 2016<br>**Data collection methods:**<br>Survey pre- and post-course; Acceptability measures mid-way | **Sample size:**<br>n = 177 across 15 cohorts<br>**Participants:**<br>88% female<br>Age (mean ± SD); 41.81 ± 11.90 years<br>Experience (mean ± SD); 4.76 ± 6.22 years<br>**Setting:**<br>Scotland<br>Healthcare (health and social care practitioners)<br>**Comparator intervention or control:**<br>N/A | **Training type/ content**<br>"Helping People Change for Health" Online module: 'health behaviour change level 1' designed by NHS Scotland.<br>Training: designed by psychologists with expertise in behavioural science.<br>**Online/ face-to-face**<br>Mixture; pre-course e-learning online module followed by face-to-face training.<br>**Training duration**<br>2 days face-to-face, held a week apart.<br>**Trainer**<br>Four health psychologists, one clinical psychologist, five health improvement specialists working in pairs. Facilitators trained by overall course lead*. | **Primary findings:**<br>*Trainee reaction to training and preferred elements:* Practice skills 'active learning' and peer to peer feedback.<br>Group facilitation- meeting others and sharing experiences and knowledge.<br>Tailored learning (e.g. case studies and examples taken from participants needs assessment).<br>Conversation frameworks, listening skills (e.g. open discovery questions (ODQs)), techniques to use.<br>Methods of teaching- e.g. seeing trainers demonstrate. | Mixed methods study, see Table 3 for full study details.<br>*Facilitator training consisted of: a) 10 hours reading, b) observation of one full course, c) two hours one-to-one discussion and practice, d) co-facilitation of parts of course plus |
| Chisholm et al. (2019)<br>https://doi.org/10.1093/pubmed/fdy094 | **Study design:**<br>Qualitative<br>**Dates of data collection:**<br>January – June 2017<br>**Data collection methods:**<br>Semi-structured interviews<br>**Outcomes of interest:** | **Sample size:**<br>n = 13<br>**Participants:**<br>Public Health practitioners<br>61.5% female<br>Age (mean ± SD); 49.5 ± 9.12 years<br>Current post average duration (mean ± SD); 5 ± 5.53 years (range 1 month to 18 years) | **Training type/ content**<br>MECC, some using existing programmes, some bespoke programmes some evaluating previous programmes<br>**Online/ face-to-face**<br>Unclear, varied between Trusts<br>**Training duration**<br>Unclear, varied between Trusts | **Primary findings:**<br>*Trainee reaction to training and preferred elements:* Broader topics means relevant to more staff, but can prevent constructive discussion with patients. Narrow scope can fit in with trainer expertise/ local priorities. Some standardisation would be good, but need to allow for local tailoring. | Self-selecting sample |
|  | Views of MECC and experience of implementing<br>**Outcome measures:**<br>Inductive thematic analysis of interview responses | **Setting:**<br>England<br>Hospital trust, community trust, local authority<br>**Comparator intervention or control:**<br>N/A | **Trainer**<br>Unclear, varied between Trusts | Online is efficient use of time and resource, but face-to-face could be needed as subject is complex.<br>Should be mandatory (but doesn't necessarily translate to use in practice).<br>Local advocates/ champions useful.<br>*Experience and perception of barriers and facilitators for MECC training and utilisation in practice:* Can be seen as an add-on to current workload. Tension between current reactive system, and health promotion.<br>Lack of supportive infrastructure, momentum, and staff engagement.<br>Commissioning difficulties and lack of resources (trainers/ money for training resources).<br>Trainer expertise/ organisation policy and priorities or lack of evidence. |  |
| Chisholm et al. (2020)<br>https://doi.org/10.1186/s12913-020-05264-9 | **Study design:**<br>Pre-post survey<br>**Dates of data collection:**<br>Not reported<br>**Data collection methods:** | **Sample size:**<br>n = 206<br>**Participants:**<br>91% female<br>Age range 22 to 62 years<br>48% participants worked in children and families service | **Training type/ content**<br>MECC behaviour change, based on 'TEnt PEGS' toolkit*. Included communication skills, understanding health behaviour and its complexities, behaviour change techniques and implementation intentions and action planning. | **Primary findings:**<br>*Trainee reaction to training and preferred elements:* Content can seem too rigid, does not reflect day-to-day practice and too lengthy | Mixed methods study, see Table 3 for full study details.<br>*TEnt PEGS toolkit was the basis for the training (Chisholm et al. 2014) |
|  | Survey immediately before and after training module (both optional) | **Setting:**<br>England<br>Mental health and community trust<br>**Comparator intervention or control:**<br>N/A | **Online/ face-to-face**<br>Online interactive module with multiple choice questions and open answer questions.<br>**Training duration**<br>40 minutes<br>**Trainer**<br>Self-led | Some would have preferred face-to-face training so could practice and have fewer distractions.<br>Learning communication skills, behaviour change techniques and patient-led approaches. |  |
| Dewhurst & Speller (2015) | **Study design:**<br>Feasibility/ pilot study<br>**Dates of data collection:**<br>Process evaluation interviews, 7 at HHFT (Nov 2013 – Mar 2014), 3 at PCC (Mar - Apr 2014), 4 at SHFT (Jun - Jul 2014)<br>**Data collection methods:**<br>Evaluation interviews with senior managers, pilot leads and front-line staff. | **Sample size:**<br>SHFT: n = 63; HHFT: n = 45; PCC: n = 15<br>**Participants:**<br>Mostly female (84% pre- and 87% post-)<br>Mostly aged 25-54 (82% pre- and 87% post-)<br>40% in role 1-5 years, 13% in role under 1 year<br>**Setting:**<br>England<br>Healthcare, Occupational Health, local area housing office (PCC)<br>**Comparator intervention or control:**<br>N/A | **Training type/ content**<br>Wessex MECC intervention and organisational changes.<br>SHFT and HHFT included all HCS competencies.<br>PCC did not include any HCS competencies.<br>**Online/ face-to-face**<br>Face-to-face, HHFT also have pre-course video<br>**Training duration**<br>‘Train the trainer’: Two 3-hour group sessions and one 6-hour group session<br>Local training:<br>SHFT 1 x 6-hour<br>HHFT 1 x 4-hour (therapy services) OR 3 x 3-hour (diabetes) OR 2 x 2-hour (health4work) session<br>PCC 1x3hr<br>**Trainer**<br>First: HCS train the trainer attended by pilot leads and some staff.<br>Second: local training delivered by leads who attended the initial training. | **Primary findings:**<br>*Trainee reaction to training and preferred elements:* Staff who are experienced trainers, prepared adequately and confident should deliver training. Mixed feedback on train-the-trainer, unsure if experienced/ knowledgeable enough.<br>Refresher sessions and support should be done regularly.<br>Could do brief introduction via e-learning (e.g. MECC info/ organisational policy), manageable chunks.<br>Include 'behaviour change' in all professional training.<br>Practice techniques, so face-to-face preferred over online.<br>Consistency good but useful to make relevant to setting as well. | Mixed methods study, see Table 3 for full study details. |
|  |  |  |  | Include non-clinical staff (i.e. those who are not providing direct patient care). |  |
| Lawrence et al. (2022)<br>https://doi.org/10.1177/1757913920977030 | **Study design:**<br>Longitudinal feasibility study<br>**Dates of data collection:**<br>Unclear.<br>**Data collection methods:**<br>Pre- and post- training evaluation.<br>Reflective feedback interviews at 1-2 months and 11-13 months post-training. | **Sample size:**<br>n = 15<br>**Participants:**<br>Age; range 20-60 years<br>Experience; mean 10 years.<br>Sex distribution unclear<br>**Setting:**<br>England<br>General practitioner practices<br>**Comparator intervention or control:**<br>N/A | **Training type/ content**<br>HCS as part of CPD<br>**Online/ face-to-face**<br>Face to face group sessions, no PowerPoint slides, interactive/ participatory. Followed by ongoing support which can be a phone call or visit by trainer to discuss how skills are being implemented (*timeframe for this unclear*)<br>**Training duration**<br>2 x 3-4 hours over 1 week<br>**Trainer**<br>Two HCS trainers plus another team member, all experienced in group work and behavior change | **Primary findings:**<br>*Trainee reaction to training and preferred elements:* Interactive nature of training/ participatory style<br>Being taught in groups with other healthcare professionals<br>Post-training feedback/ follow-up useful<br>*Experience and perception of barriers for MECC training and utilisation in practice:* Patients medical conditions/ habitual nature/ circumstances or general attitude<br>Time pressures, expectation to signpost | Mixed methods study, see Table 3 for full study details. |
| Mills et al. (2021)<br>https://doi.org/10.12968/bjcn.2021.26.11.554 | **Study design:**<br>Survey<br>**Dates of data collection:**<br>Not reported<br>**Data collection methods:**<br>Online questionnaire | **Sample size:**<br>n = 108<br>**Participants:**<br>Student nurses<br>Age/ sex distribution unclear<br>**Setting:**<br>England<br>Adult nursing<br>**Comparator intervention or control:**<br>N/A | **Training**<br>MECC HCS<br>**Online/ face-to-face**<br>Face-to-face as part of health promotion module of second year of nursing degree<br>**Training duration**<br>Unclear<br>**Trainer**<br>Nurse academics who deliver the module. Had undergone a MECC trainer's course. | **Primary findings:**<br>*Experience and perception of barriers and facilitators for MECC training and utilisation in practice:* Lack of time.<br>Lack of knowledge of local services.<br>Lack of mentor knowledge/ role modelling of HCS. | Mixed methods study, see Table 3 for full study details. |
| Nelson et al. (2013)<br>http://dx.doi.org/10.1016/j.puhe.2013.04.013 | **Study design:**<br>Semi-structured interview<br>**Dates of data collection:**<br>Aug - Sept 2011<br>**Data collection methods:**<br>Semi structured face-to-face or telephone interviews.<br>**Outcomes of interest:**<br>Evaluation of MECC, including questions related to training and development, implementation and delivery, impact, and sustainability.<br>**Outcome measures:**<br>Thematic analysis. | **Sample size:**<br>n = 12, professionals involved in design, delivery, or evaluation of MECC<br>**Participants:**<br>Varying roles in Public Health, Primary Care Trusts, Private Sector and Local Authority. Age, sex, and experience distribution unclear.<br>**Setting:**<br>England<br>NHS/ local authority/ private sector<br>**Comparator intervention or control:**<br>N/A | **Training type/ content**<br>MECC training/ PLBC framework which is split into four competency levels*<br>Encourages signposting to other services.<br>**Online/ face-to-face**<br>Unclear, but appeared to be face-to-face.<br>**Training duration**<br>Level 1 training is a half day, unclear whether the other levels can be combined, or how long they take.<br>**Trainer**<br>Unclear, but as study interviews professionals involved in delivery of MECC, likely to be specialist MECC trainers. | **Primary findings:**<br>*Trainee reaction to training and preferred elements:*<br>Accreditation (CPD)/ recognition in PDRs.<br>Train the trainer effective (critical mass/ saturation).<br>Focus on improving skills and confidence, not too many facts/ background.<br>Tailor to fit to services and clients' needs (collaborative approach- awareness of practitioner issues and service user issues).<br>Low cost and low time (1/2 day for level 1).<br>Train non-medical staff e.g. porters/ receptionists.<br>Continual follow-up/ support services to refer people too.<br>*Experience and perception of barriers and facilitators for MECC training and utilisation in practice:* Asking staff to do more.<br>Concern of offending clients.<br>Other time pressures. | Interview questions included asking about training conducted and what worked well or could be improved, success factors or barriers to implementing MECC and lessons learnt, staff feedback about MECC and any factors that can influence the spread of MECC.<br>*Level 1: introduce idea of lifestyle behaviour change and motivate individuals to consider/ think about making changes.<br>Level 2: use brief behaviour change techniques (BCTs) to help individuals take action.<br>Level 3: Provide support as individuals change lifestyle behaviours and facilitate them to maintain these changes.<br>Level 4: Use specialist BCTs to support individuals where brief interventions have not been successful or who have more complex needs. |
| Pallin et al. (2022) | **Study design:**<br>Sequential mixed method.<br>**Dates of data collection:** | **Sample size:**<br>n = 37; 16 completed survey after nutrition course, 21 after | **Training type/ content**<br>Courses on engaging in healthy diet and physical activity conversations | **Primary findings:**<br>*Trainee reaction to training and preferred elements:* Mandatory | Mixed methods study, see Table 3 for full study details including course names. |
| https://doi.org/10.1016/j.radi.2021.09.004 | May 2020 to July 2020<br>**Data collection methods:**<br>Online survey/ telephone interview. Participants were asked to complete the survey immediately after completing the training. | physical activity course. 21 completed telephone interviews.<br>**Participants:**<br>Therapeutic radiographer (no other data given)<br>**Setting:**<br>England<br>Cancer healthcare<br>**Comparator intervention or control:**<br>N/A | with people living with and beyond cancer.<br>**Online/ face-to-face**<br>All online- mixed of courses (video/ interactive) and websites.<br>**Training duration**<br>One of the courses was 30 minutes, others unclear duration. Participants were randomised to undertake 2 of 5 identified relevant courses.<br>**Trainer**<br>Unclear, but all appear to be self-led. | training increases confidence in training.<br>Include with induction training/ protected CPD time.<br>Online courses can develop knowledge of why the delivery of advice is important, and how to deliver advice.<br>Important to give evidence and scientific rationale, e.g. stats on benefits of exercise/ healthy eating, physiology of it.<br>Need to see relevance of course to their role and patient needs (i.e. ensure targeted to population the practitioners will see and the specific issues they may face).<br>Need managerial support.<br>*Experience and perception of barriers and facilitators for MECC training and utilisation in practice:* Lack of clear guidance and clarity around professional boundaries.<br>Organisational commitment/ leadership. |  |
| Parchment et al. (2023)<br>https://doi.org/10.1186/s12913-023-10126-1 | **Study design:**<br>Mixed methods, sequential explanatory study<br>**Dates of data collection:**<br>Nov 2020-Apr 2021 | **Sample size:**<br>n = 11<br>**Participants:**<br>Age; mean 41.8 (SD 8.4) years; range 29 to 53 years. | **Training type/ content**<br>All did HCS Lite<br>Two did full MECC training<br>Three did HCS train-the-trainer programme. | **Primary findings:**<br>*Trainee reaction to training and preferred elements:* Refreshers/ reminders annually. | This paper reports the qualitative findings of this mixed methods study. A previous publication reported quantitative results, but |
|  | **Data collection methods:**<br>Online survey<br>**Outcomes of interest:**<br>Gain an in-depth, qualitative understanding of the views, experiences, and acceptability of MECC HCS for the physiotherapists. Discuss qualitative findings in relation to behaviour change theory to provide recommendations for future implementation.<br>**Outcome measures:**<br>Semi-structured interview; exploratory, open questions to gain a deeper understanding of the physiotherapists' experiences and perceived acceptability of implementing MECC HCS in routine practice. | 82% female.<br>**Setting:**<br>Physiotherapists in the UK working with patients living with MSK conditions and/or pain.<br>**Comparator intervention or control:**<br>N/A | **Online/ face-to-face**<br>Unclear.<br>**Training duration**<br>HCS Lite: 1x 3-hour<br>MECC HCS: 2 x 3-hour<br>Train-the-trainer: Unclear duration<br>**Trainer**<br>Unclear. | Offer at beginning of career (e.g. in undergraduate programmes).<br>Practice in training/ real life.<br>Having a local lead/ champion to ensure MECC is sustained and training more accessible.<br>*Experience and perception of barriers and facilitators for MECC training and utilisation in practice:* Time in routine appointments to deliver intervention.<br>Time to attend formal training to develop skills.<br>Resistance from management to allow staff to attend training. | these did not cover MECC training.<br>All participants had to be using HCS in practice daily, and have attended training at least one year ago. |
| Tuohy et al. (2021)<br>https://doi.org/10.1016/j.nepr.2021.103246 | **Study design:**<br>Descriptive qualitative study.<br>**Dates of data collection:**<br>Module taught in Spring 2019<br>**Data collection methods:**<br>Focus group interviewing.<br>**Outcomes of interest:** | **Sample size:**<br>n = 17 (out of 131 students)<br>**Participants:**<br>16 nursing, 1 midwifery<br>88% female<br>29% mature students (23 years and over), 71% school leaver (18 to 22 years)<br>**Setting:**<br>Ireland | **Training type/ content**<br>MECC units one and two* included in first year health promotion module in BSc nursing and midwifery programmes.<br>Units three and four taught in third and fourth years, were not evaluated in the study and therefore not explained further here.<br>**Online/ face-to-face** | **Primary findings:**<br>*Trainee reaction to training and preferred elements:* Linked with anatomy/ physiology psychology so shows why it is important to patients and self.<br>Link between health determinants and people's behaviour.<br>Different people need different things- some like to read, some | *MECC unit 1: Health and personal wellness, includes biopsychosocial model of health and lifestyle influences on health.<br>MECC unit 2: Lifestyle behaviours and personal responsibility for health, includes theories of behaviour changed. |
|  | Students' experiences of the teaching and learning of MECC in their undergraduate nursing and midwifery training and education<br>**Outcome measures:**<br>Inductive thematic analysis | Healthcare training and education<br>**Comparator intervention or control:**<br>N/A | Face-to-face including lectures, group work, case studies. Some self-led reading also required.<br>**Training duration**<br>Unclear what proportion of the module time was MECC units one and two training.<br>**Trainer**<br>Unclear, but there were 'visiting speakers' in addition to lecturers. | like practical exercises- allow for this in training.<br>Being taught techniques on enabling patients to think about their own health behaviours.<br>Liked being in groups with others- exposure to difference perspectives.<br>Variety of speakers and teaching strategies. |  |

**Abbreviations:** BCT: Behaviour change techniques, CPD: Continued professional development, HCS: Healthy Conversation Skills, HHFT: Hampshire Hospitals NHS Foundation Trust, MECC: Making Every Contact Count, N/A: Not applicable, NHS: National Health Service, ODQ: open discovery questions, PCC: Portsmouth City Council, PDR: Personal development review, PLBC: Prevention and Lifestyle Behaviour Change: Competence Framework, SD: Standard deviation, SHFT: Southern Health NHS Foundation Trust, TeNT PEGS: T = Taking down barriers; EN = Changing the ENvironment; Th = Addressing Thoughts and emotions; P = Perform and practice; E = Empowering people to change; G = Achieving Goals; S = Social support.

### 6.3 Quality appraisal

**Table 5:**
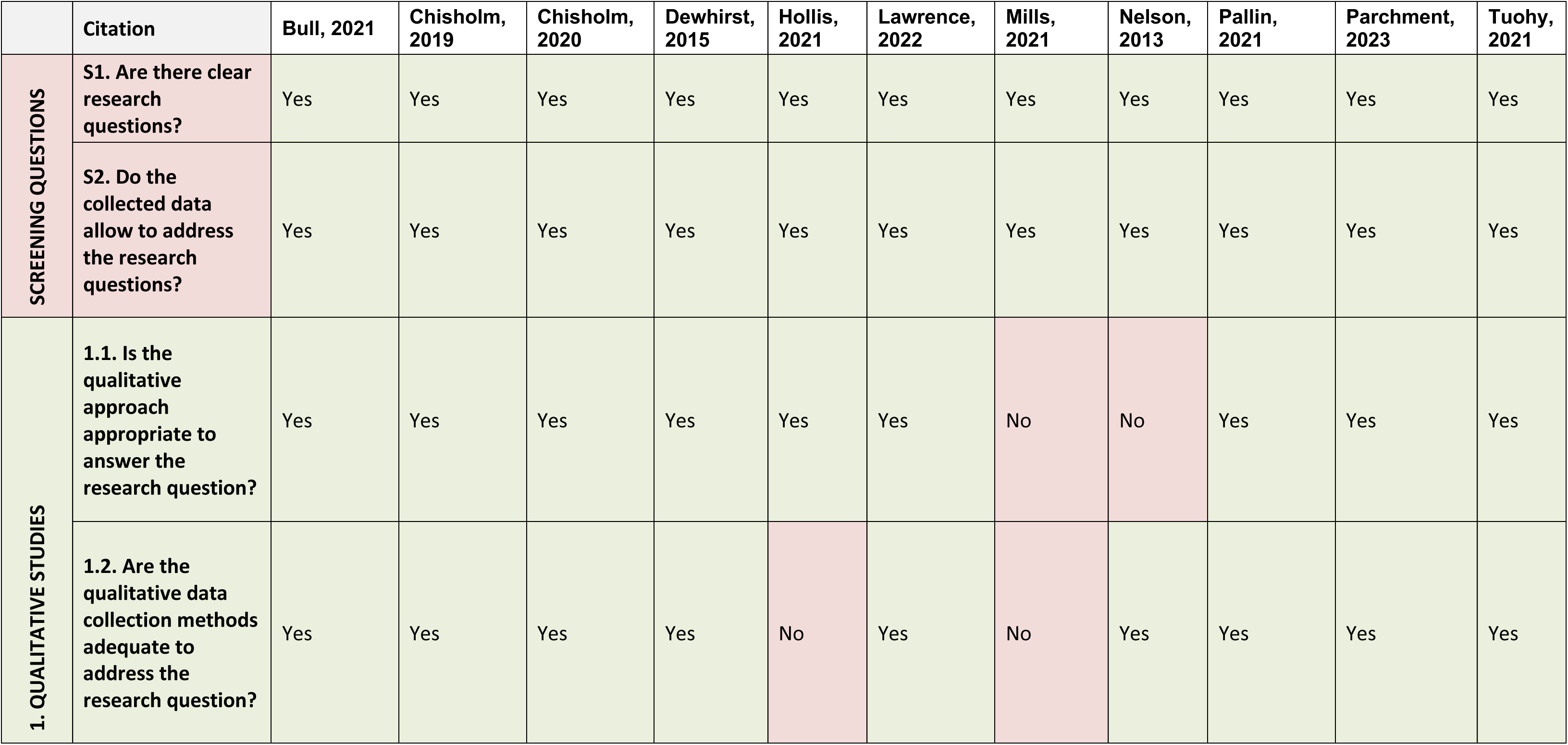
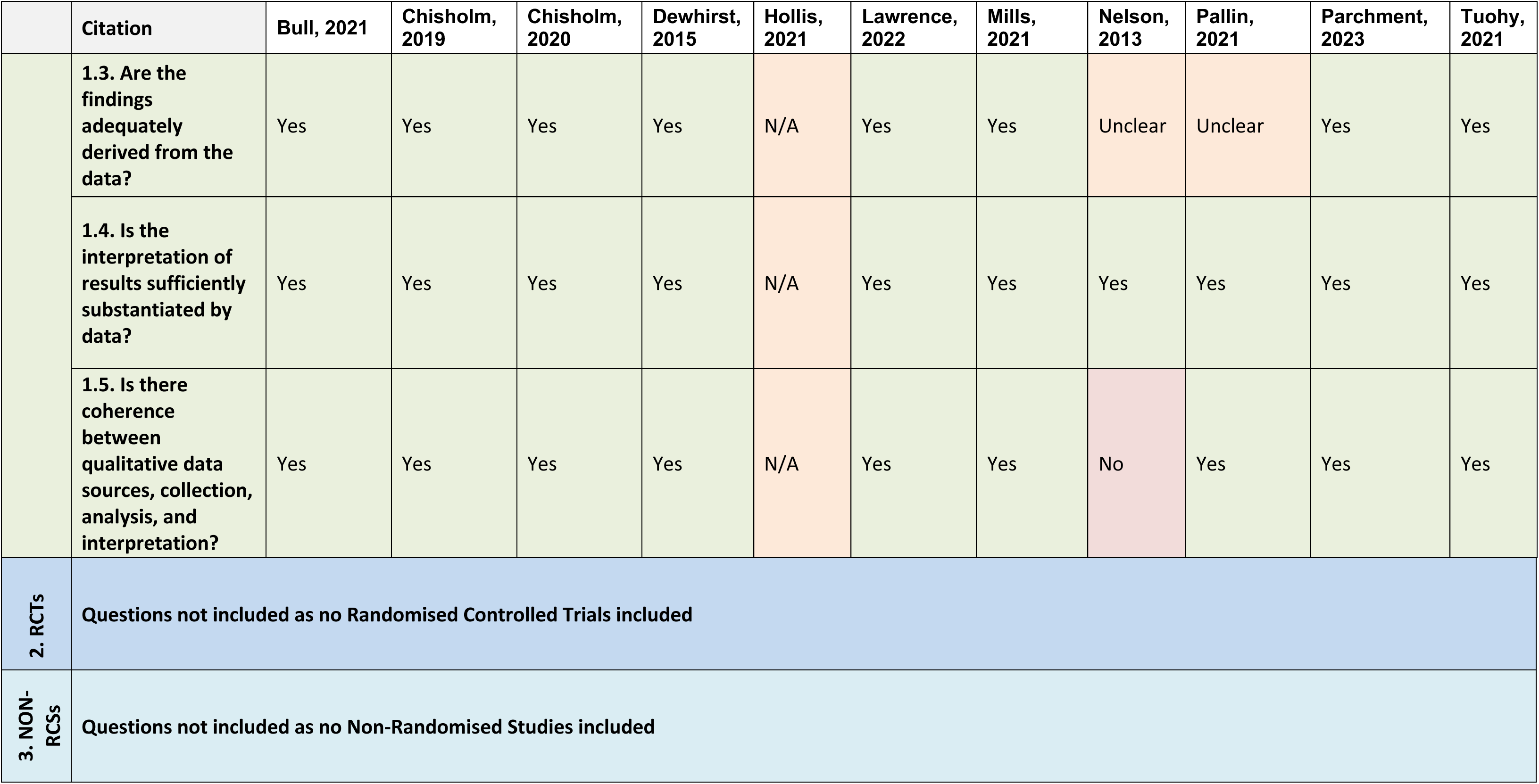
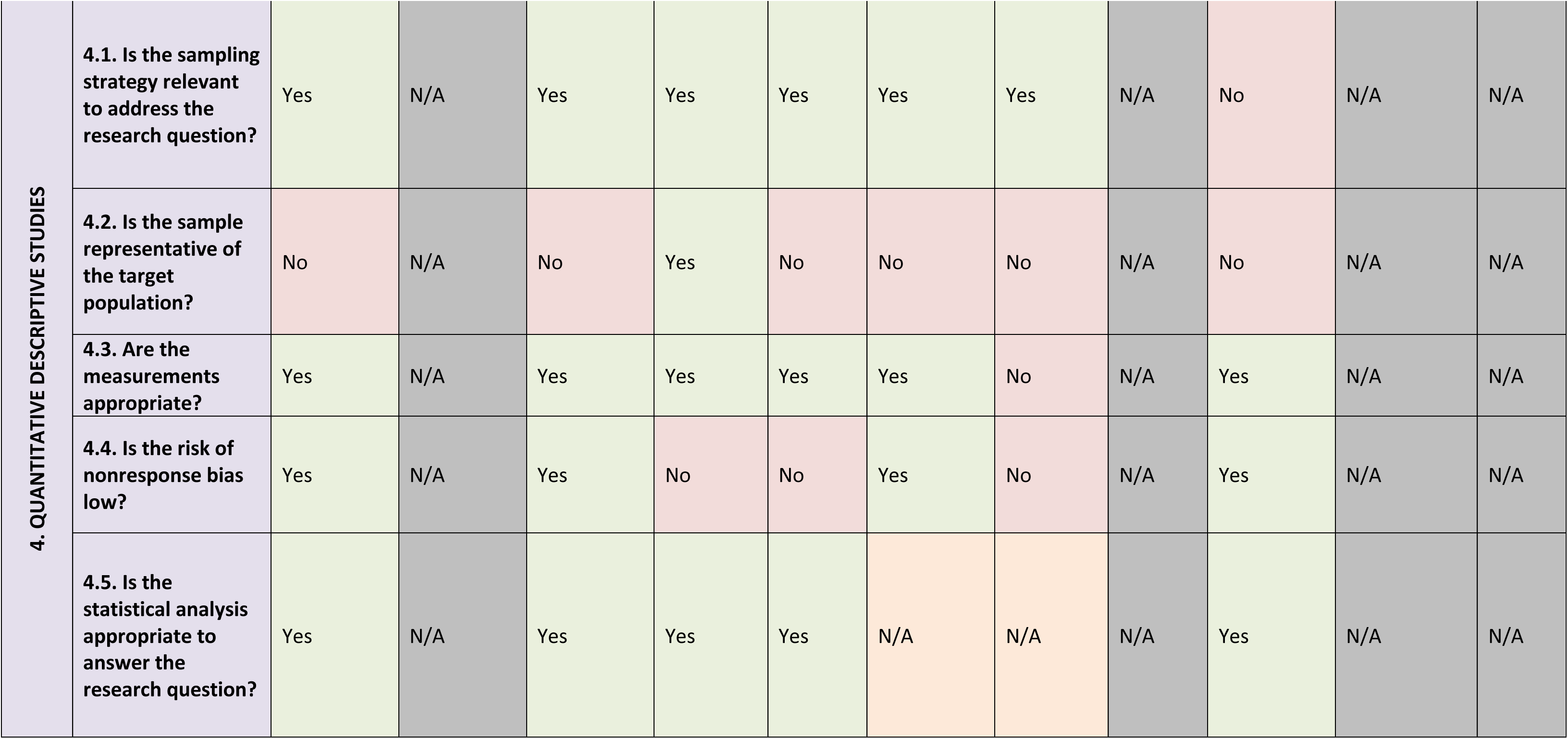
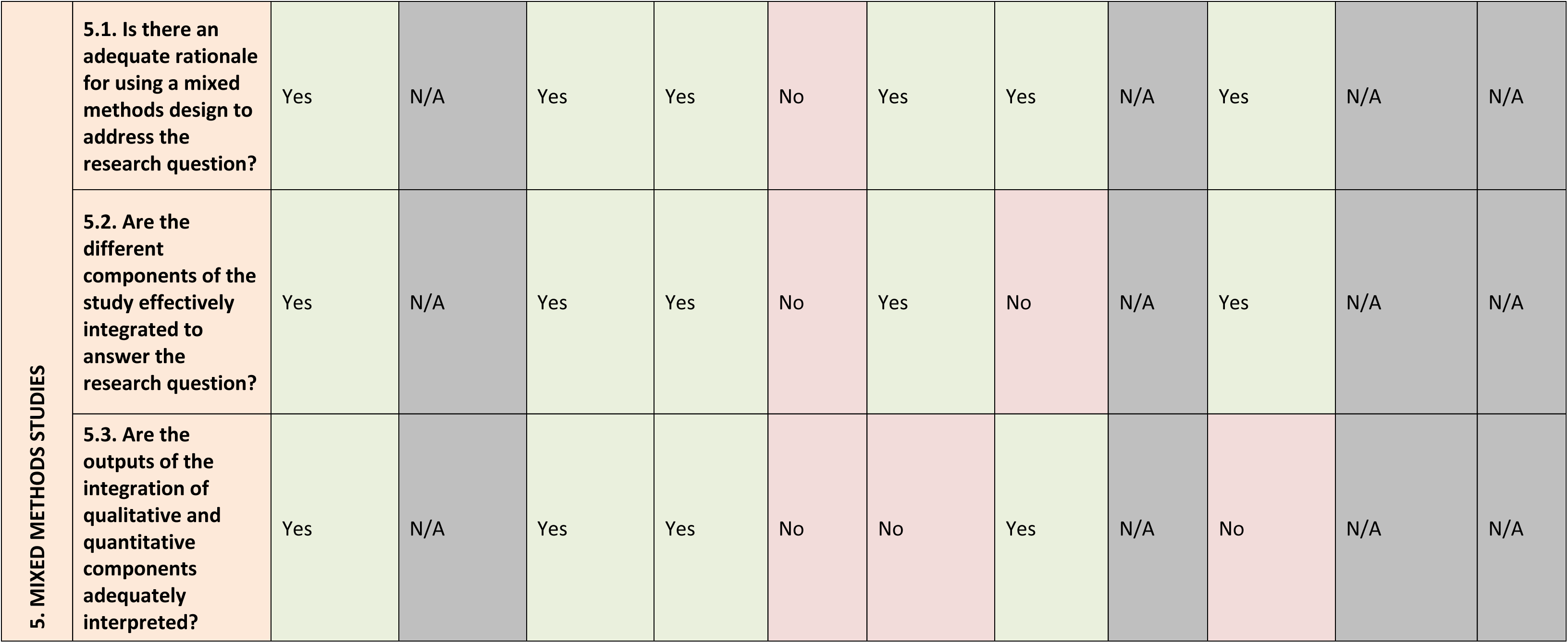
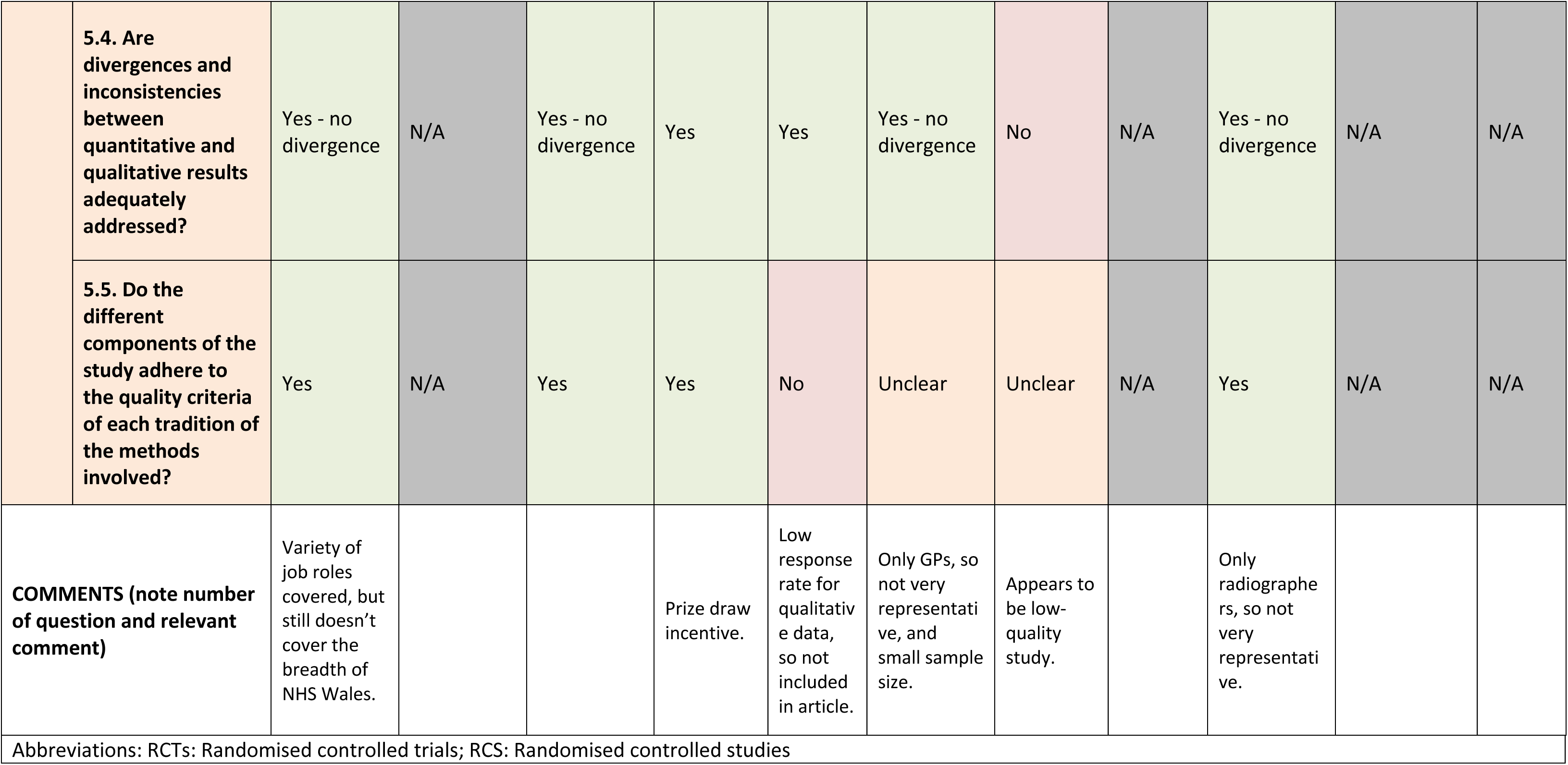
Completed Mixed Methods Appraisal Tool.

|  | Citation | Bull, 2021 | Chisholm, 2019 | Chisholm, 2020 | Dewhurst, 2015 | Hollis, 2021 | Lawrence, 2022 | Mills, 2021 | Nelson, 2013 | Pallin, 2021 | Parchment, 2023 | Tuohy, 2021 |
| --- | --- | --- | --- | --- | --- | --- | --- | --- | --- | --- | --- | --- |
| SCREENING QUESTIONS | S1. Are there clear research questions? | Yes | Yes | Yes | Yes | Yes | Yes | Yes | Yes | Yes | Yes | Yes |
|  | S2. Do the collected data allow to address the research questions? | Yes | Yes | Yes | Yes | Yes | Yes | Yes | Yes | Yes | Yes | Yes |
| 1. QUALITATIVE STUDIES | 1.1. Is the qualitative approach appropriate to answer the research question? | Yes | Yes | Yes | Yes | Yes | Yes | No | No | Yes | Yes | Yes |
|  | 1.2. Are the qualitative data collection methods adequate to address the research question? | Yes | Yes | Yes | Yes | No | Yes | No | Yes | Yes | Yes | Yes |
|  | 1.3. Are the findings adequately derived from the data? | Yes | Yes | Yes | Yes | N/A | Yes | Yes | Unclear | Unclear | Yes | Yes |
|  | 1.4. Is the interpretation of results sufficiently substantiated by data? | Yes | Yes | Yes | Yes | N/A | Yes | Yes | Yes | Yes | Yes | Yes |
|  | 1.5. Is there coherence between qualitative data sources, collection, analysis, and interpretation? | Yes | Yes | Yes | Yes | N/A | Yes | Yes | No | Yes | Yes | Yes |
| 2. RCTs | Questions not included as no Randomised Controlled Trials included |  |  |  |  |  |  |  |  |  |  |  |
| 3. NON-RCTs | Questions not included as no Non-Randomised Studies included |  |  |  |  |  |  |  |  |  |  |  |
| 4. QUANTITATIVE DESCRIPTIVE STUDIES | 4.1. Is the sampling strategy relevant to address the research question? | Yes | N/A | Yes | Yes | Yes | Yes | Yes | N/A | No | N/A | N/A |
|  | 4.2. Is the sample representative of the target population? | No | N/A | No | Yes | No | No | No | N/A | No | N/A | N/A |
|  | 4.3. Are the measurements appropriate? | Yes | N/A | Yes | Yes | Yes | Yes | No | N/A | Yes | N/A | N/A |
|  | 4.4. Is the risk of nonresponse bias low? | Yes | N/A | Yes | No | No | Yes | No | N/A | Yes | N/A | N/A |
|  | 4.5. Is the statistical analysis appropriate to answer the research question? | Yes | N/A | Yes | Yes | Yes | N/A | N/A | N/A | Yes | N/A | N/A |
| 5. MIXED METHODS STUDIES | 5.1. Is there an adequate rationale for using a mixed methods design to address the research question? | Yes | N/A | Yes | Yes | No | Yes | Yes | N/A | Yes | N/A | N/A |
|  | 5.2. Are the different components of the study effectively integrated to answer the research question? | Yes | N/A | Yes | Yes | No | Yes | No | N/A | Yes | N/A | N/A |
|  | 5.3. Are the outputs of the integration of qualitative and quantitative components adequately interpreted? | Yes | N/A | Yes | Yes | No | No | Yes | N/A | No | N/A | N/A |
|  | 5.4. Are divergences and inconsistencies between quantitative and qualitative results adequately addressed? | Yes - no divergence | N/A | Yes - no divergence | Yes | Yes | Yes - no divergence | No | N/A | Yes - no divergence | N/A | N/A |
|  | 5.5. Do the different components of the study adhere to the quality criteria of each tradition of the methods involved? | Yes | N/A | Yes | Yes | No | Unclear | Unclear | N/A | Yes | N/A | N/A |
| **COMMENTS (note number of question and relevant comment)** |  | Variety of job roles covered, but still doesn't cover the breadth of NHS Wales. |  |  | Prize draw incentive. | Low response rate for qualitative data, so not included in article. | Only GPs, so not very representative, and small sample size. | Appears to be low-quality study. |  | Only radiographers, so not very representative. |  |  |
| Abbreviations: RCTs: Randomised controlled trials; RCS: Randomised controlled studies |  |  |  |  |  |  |  |  |  |  |  |  |

### 6.4 Information available on request

Protocol and excluded studies.

## 7.2 Acknowledgements

The authors would like to thank Rhian Meaden, Charlotte Cullum, Megan Elliott and Nathan Davies for their time, expertise, and contributions, including in guiding the focus of the review and interpretation of findings.

## 8. APPENDIX: Search strategies

### Database

Ovid MEDLINE(R) ALL <1946 to June 25, 2024>

| # | Query | Results from 26 Jun 2024 |
| --- | --- | --- |
| 1 | "making every contact count".tw. | 49 |
| 2 | "every day interaction*".tw. | 3 |
| 3 | "healthy conversation*".tw. | 18 |
| 4 | (brief adj1 (intervention* or interaction* or conversation*)).tw. | 5,356 |
| 5 | "behavio?r change".tw. | 21,115 |
| 6 | 4 and 5 | 196 |
| 7 | 1 or 2 or 3 or 6 | **255** |

### Database

Embase Classic+Embase <1947 to 2024 June 25>

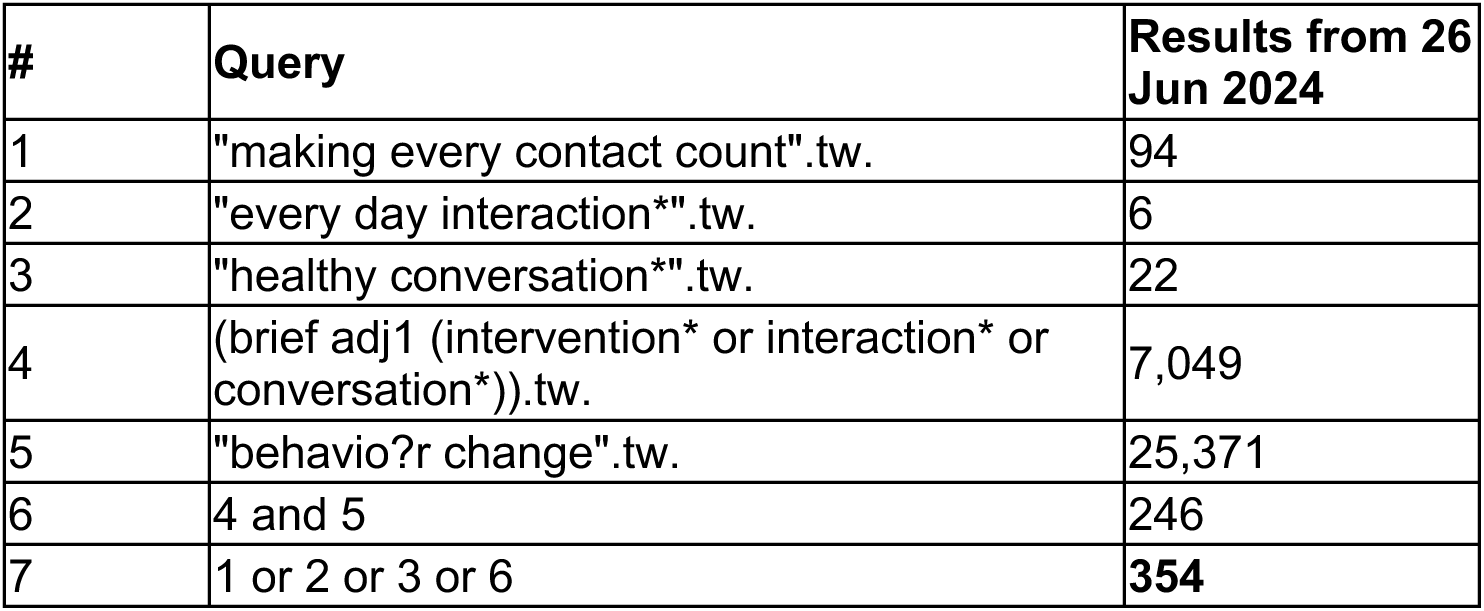

### Database

CINAHL: 27^th^ June 2024

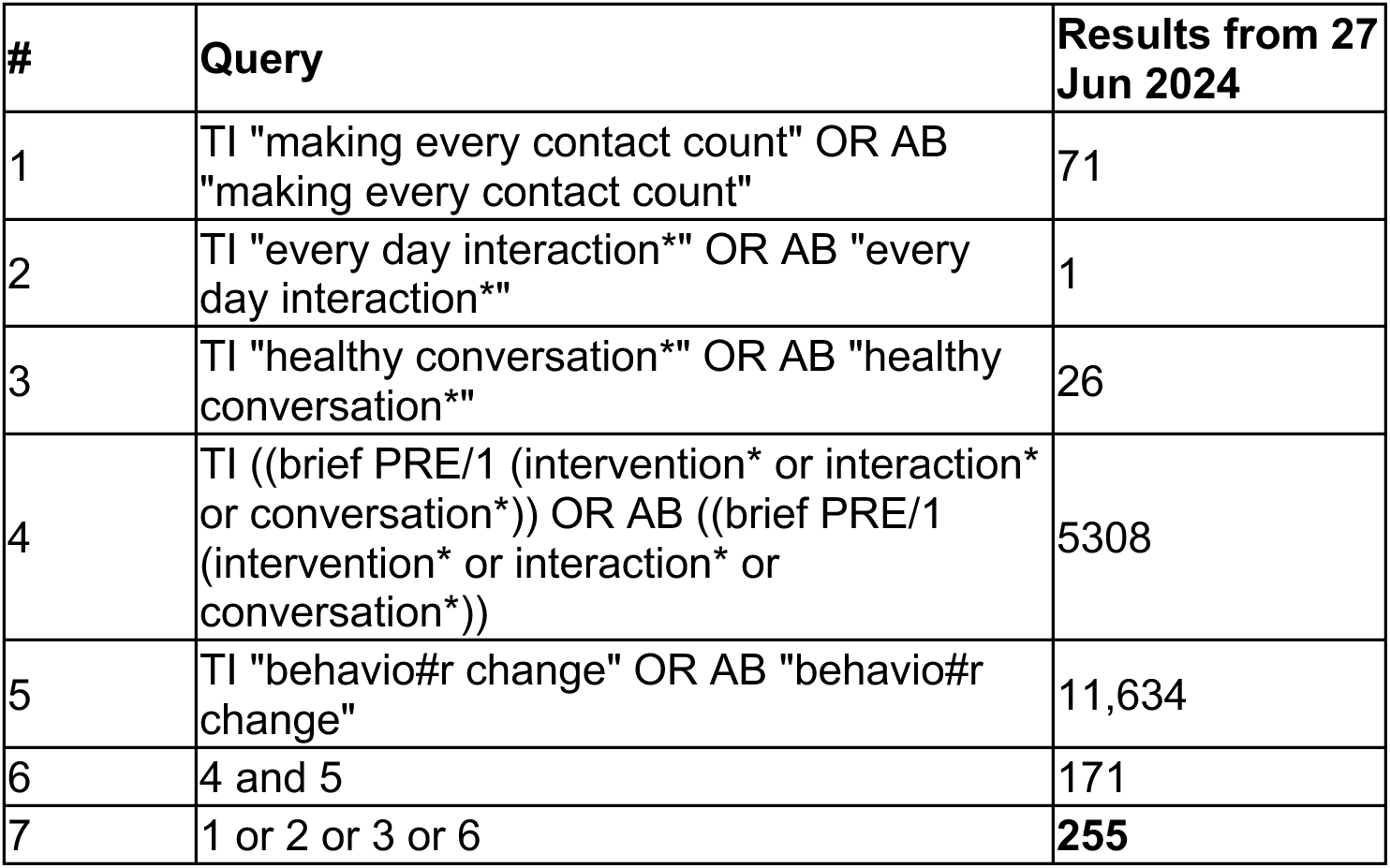

### Database

Scopus: 27^th^ June 2024

**Database:**Scopus: 27<sup>th</sup> June 2024
| # | Query | Results from 27 Jun 2024 |
| --- | --- | --- |
| 1 | TITLE-ABS-KEY ("making every contact count") | 66 |
| 2 | TITLE-ABS-KEY ("every day interaction*") | 23 |
| 3 | TITLE-ABS-KEY ("healthy conversation*") | 49 |
| 4 | TITLE-ABS-KEY (brief PRE/1 (intervention* or interaction* or conversation*)) | 10,423 |
| 5 | TITLE-ABS-KEY ("behavio?r change") | 76,518 |
| 6 | 4 and 5 | 600 |
| 7 | 1 or 2 or 3 or 6 | **721** |

### Database

Web of Science: 27^th^ June 2024

**Database:**Web of Science: 27<sup>th</sup> June 2024
| # | Query | Results from 27 Jun 2024 |
| --- | --- | --- |
| 1 | "making every contact count" (Topic) | 58 |
| 2 | "every day interaction*" (Topic) | 4 |
| 3 | "healthy conversation*" (Topic) | 15 |
| 4 | brief NEAR/1 (intervention* or interaction* or conversation*) (Topic) | 11,969 |
| 5 | "behavio?r change" (Topic) | 40,423 |
| 6 | 4 and 5 | 508 |
| 7 | 1 or 2 or 3 or 6 | **605** |

### Overton

27^th^ June 2024

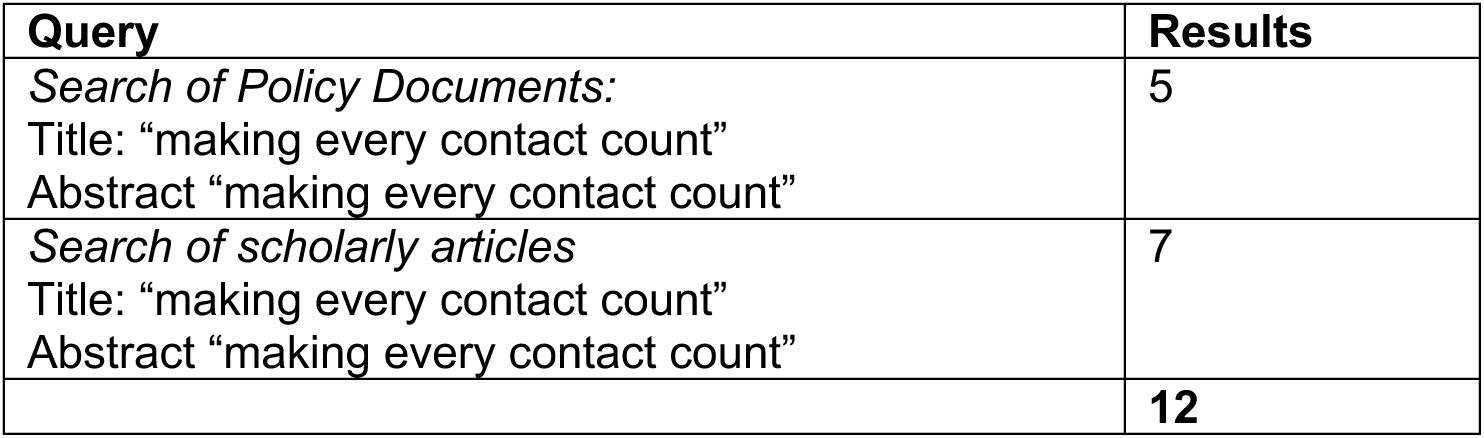

**Overton: 27<sup>th</sup> June 2024**
| Query | Results |
| --- | --- |
| *Search of Policy Documents:*<br>Title: "making every contact count"<br>Abstract "making every contact count" | 5 |
| *Search of scholarly articles*<br>Title: "making every contact count"<br>Abstract "making every contact count" | 7 |
|  | **12** |

### Total Numbers retrieved

|  |  |
| --- | --- |
| Medline (Ovid) | 255 |
| EMBASE (Ovid) | 354 |
| CINAHL (EBSCO) | 255 |
| Scopus | 721 |
| Web of Science | 605 |
| Overton | 12 |
| Total | **2202** |
| Duplicates removed in Endnote | 652 |
| Total | **1550** |

## Footnotes

* This section has been completed by the Centre for Health Economics & Medicines Evaluation (CHEME), Bangor University

